# A scoping review of non-pharmacological interventions for managing fatigue across the lifespan of people living with chronic musculoskeletal conditions

**DOI:** 10.1101/2025.07.29.25332314

**Authors:** Katie Fishpool, George Young, Coziana Ciurtin, Fiona Cramp, Emmanuel Erhieyovwe, Bayram Farisogullari, Bethan Jones, Gary Macfarlane, Pedro M. Machado, Jen Pearson, Eduardo Santos, Emma Dures

## Abstract

**Background:** Fatigue is an important and distressing symptom for many people living with chronic musculoskeletal conditions. Many non-pharmacological interventions have been investigated, and some shown to be effective in reducing fatigue, but the evidence is disparate across conditions and the lifespan.

**Objectives:** To create an overview of current knowledge by identifying existing non-pharmacological interventions for MSK fatigue across the lifespan, including their theoretical basis, characteristics of participants and the clinical competencies of those delivering interventions; to highlight gaps and collaborate with Patient and Public Involvement groups to identify priorities.

**Methods:** This scoping review followed the Joanna Briggs Institute methodology, including evidence relating to people of all ages who have been offered a non-pharmacological intervention with either the intention or effect of reducing MSK fatigue and its impact. Databases were searched for peer-reviewed primary research studies published after 1^st^ January 2007 in English language.

**Results:** Two hundred and sixty-eight eligible studies were found, including 72 studies primarily focused on fatigue. Conditions most frequently studied included fibromyalgia, rheumatoid arthritis and systemic lupus erythematous. Physical activity interventions were the most studied, most participants were adults and approximately 75% were female. Common exclusion criteria were physical comorbidities, psychiatric disease or unstable health conditions and cognitive impairments.

**Conclusions:** Few studies explore how interventions can be combined to achieve person-centred fatigue management, and many groups are routinely excluded from participating in relevant research. To effectively tailor interventions to the needs of individuals it is important to understand how their characteristics may interact with their health needs.

## INTRODUCTION

### Rationale

Musculoskeletal (MSK) conditions can be inflammatory or non-inflammatory. Conditions include connective tissue diseases, inflammatory arthritis and osteoarthritis (OA), back and neck pain, and fibromyalgia; affecting the muscles, bones, joints and connective tissue (1,2). Currently more than 20 million people in the UK and 1.7 billion people globally live with an MSK condition (1,2). Prevalence increases with age but these conditions occur across the lifespan, with approximately 234,000 children in England and Scotland living with a chronic MSK condition (2).

Fatigue has been identified by people living with chronic MSK conditions as a priority symptom which has a significant impact on quality of life (2–8). Pharmacological treatments are not licensed for the management of fatigue without concurrent disease activity, so the focus in clinical practice has been on non-pharmacological options (9,10). This has been mirrored in healthcare research and recent systematic reviews have examined the strength of the evidence supporting a range of non-pharmacological interventions in patient with a range of MSK conditions (11–15). Non-pharmacological interventions are any non-chemical or biological interventions that are theoretically based and empirically proven, or that have a logical rationale which is possible to prove by empirical study (11,16).

Recent and ongoing studies on the topic of fatigue support the need for this scoping review and have been used to inform its design. A previous scoping review explored reviews of fatigue in adults with rheumatoid arthritis (RA), OA, spondylarthritis, and fibromyalgia (17), reporting on 39 reviews of non-pharmacological interventions, of which 18 addressed fatigue as the primary objective. The authors identified several factors associated with fatigue and a range of interventions, although this was limited to published reviews and therefore did not include all study designs.

The European Alliance of Associations for Rheumatology (EULAR) recently funded a taskforce to examine the evidence for non-pharmacological interventions for fatigue in inflammatory rheumatic conditions. The resulting systematic review found that some interventions such as physical activity and psychoeducational interventions are efficacious (14). This informed the EULAR recommendations for the management of fatigue (18) and highlighted a need to better understand contextual factors and the mechanisms by which interventions are effective (7).

There is currently no comprehensive understanding of the factors which influence the success of an intervention in reducing MSK fatigue. The consequence of this is a lack of evidence to support decision making in how to design, offer and deliver interventions in the most effective way, tailored to a range of patients and at the optimal time. Current clinical guidelines for the management of common MSK conditions recognise fatigue as an important symptom but do not make any recommendations for how it can be addressed directly (19–21), hindering implementation of the evidence.

### Review aims and objectives

The aims of this scoping review were to generate evidence to inform tailored MSK-fatigue support, and to highlight gaps in the evidence base. The review objectives were to explore the:

1. Evidence for existing non-pharmacological interventions for MSK fatigue.
2. Characteristics of included participants and identify who was missing.
3. Theoretical basis for existing non-pharmacological interventions.
4. Design and outcome measures of the included studies.
5. Location of existing non-pharmacological interventions and the training and skills of those who delivered them.

### Patient and public involvement statement

Patient and Public Involvement and Engagement (PPIE) is a key aspect in designing research for real-world impact (22). The purpose of a scoping review is to understand the broad overview of the existing evidence, including what is missing. PPIE is a vital part of this process as their insights and priorities help to make sense of large heterogenous data sets and the how the evidence relates to their lived experience (23).

We have reported the outcome of PPIE events and how they influenced the scoping review process following the guidelines for the GRIPP2 short form reporting checklist (24), a tool designed to improve the reporting of public and patient involvement in research. (Appendix 1)

## METHODS

A review protocol was established in accordance with the Joanna Briggs Institute (JBI) methodology for scoping reviews (25), supported by two online PPIE workshops. These lasted for approximately one hour, with discussion facilitated by two researchers (GY and KF) and focussing on people’s experiences of offering or being offered fatigue management support. The participants attending the first workshop included adults living with one or more MSK conditions (n=6) and clinicians from a range of professions who support patients that experience MSK related fatigue (n=3). The second online workshop was attended by children and young people living with MSK conditions (n=12).

Both groups were presented with the proposed review questions, list of MSK conditions and search terms related to potential intervention types. The participants highlighted additional intervention types and pathways that were subsequently included in the search terms (Appendix 2). They also deepened the review team’s knowledge of the personal impact of experiencing MSK fatigue, confirming it as a significant issue and that an overview of potential interventions and management techniques would be welcomed by patients and clinicians.

### Systematic Literature Search

A systematic search of the literature was performed in November 2023 and citation searching was completed in February 2024, following the published review protocol (26).

**Table 1.** Eligibility criteria.

| Inclusion criteria | Exclusion criteria |
| --- | --- |
| - Primary research study<br>- Published in a peer reviewed journal<br>- Available in English language<br>- Participants have one or more chronic MSK conditions<br>- Participants experience fatigue at baseline<br>- Published during or after 2007<br>- Describes a non-pharmacological intervention to manage MSK condition symptoms, with fatigue reduction as a primary or secondary outcome | - Reviews, protocols, opinion pieces, editorials, case series, case reports, observational cohort studies<br>- Pharmacological interventions<br>- No intervention is described<br>- Muscle fatigue rather than global fatigue is examined<br>- No data is available on factors associated with intervention success (theoretical mechanism of intervention OR characteristics of participants OR characteristics of practitioners delivering interventions) |

### Study selection

Records were imported into Covidence© and duplicates were removed. Each abstract was screened by two or more independent reviewers (KF, GY, BC and EE,) and compared against the eligibility criteria to determine inclusion. Discrepancies were checked and resolved by discussion (KF, GY, ED). At full text screening stage all papers were screened by two independent reviewers (KF, GY).

The following amendments were made to the protocol during the screening process:

– The definition of fatigue as a primary outcome was reviewed by the study team to ensure consistency. Due to the heterogeneity of included studies, fatigue was considered the primary focus if this was explicitly stated, if a fatigue measurement tool had been used to determine sample size or if a hypothesis for how the intervention would act on fatigue was provided. The rationale for this was to identify the understanding and conceptual models of MSK fatigue and how these were addressed in the research, including a range of approaches to fatigue management.
– It was agreed that where multiple papers referred to the same study these would be grouped together into a single record to remove duplication while ensuring that any rich contextual data were captured.

The screening process is reported using a PRISMA flow diagram (Figure 1), including a tabulated summary of reasons for exclusion at full text screening.

**Figure 1.**
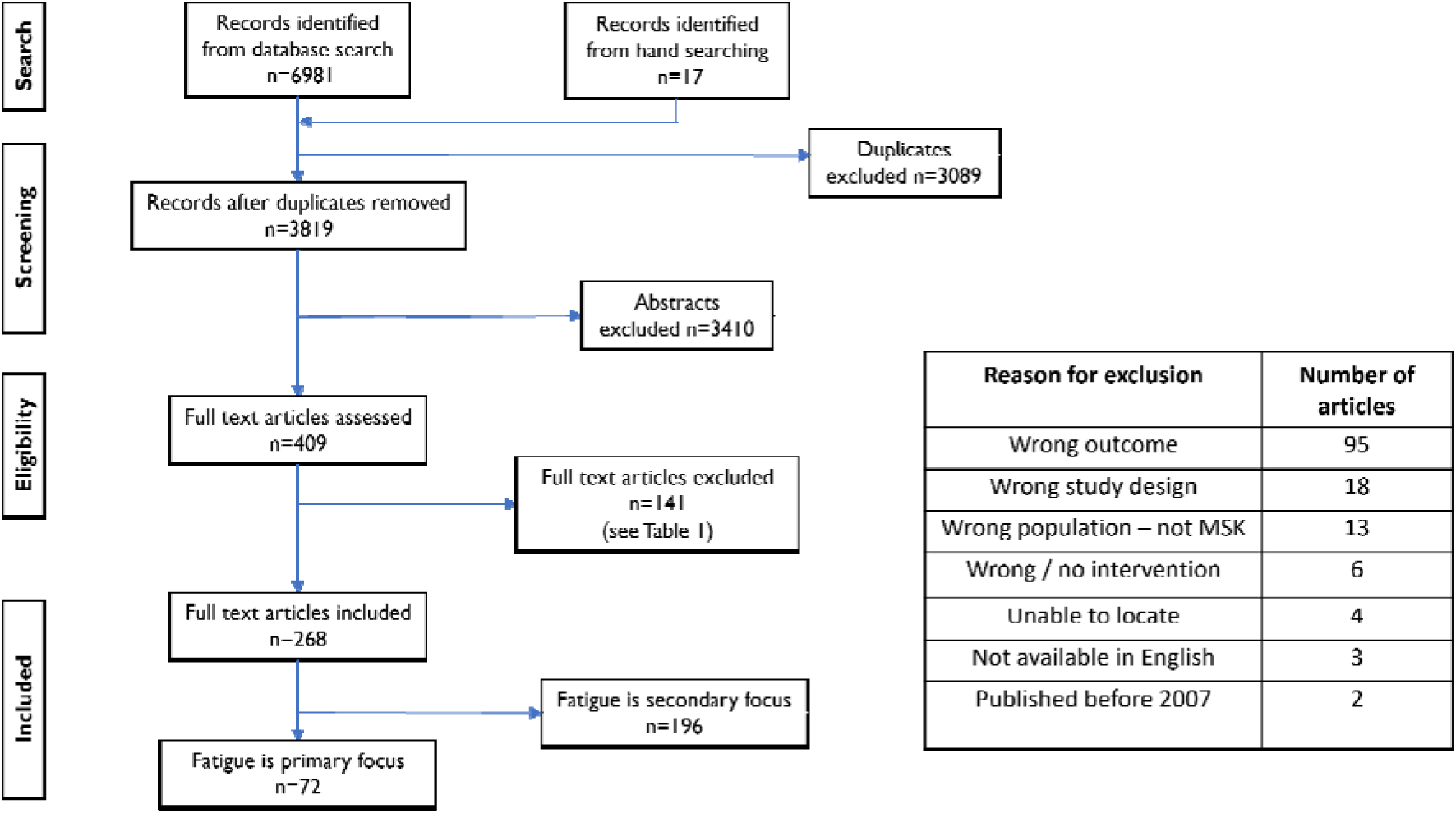

**Figure 2.**
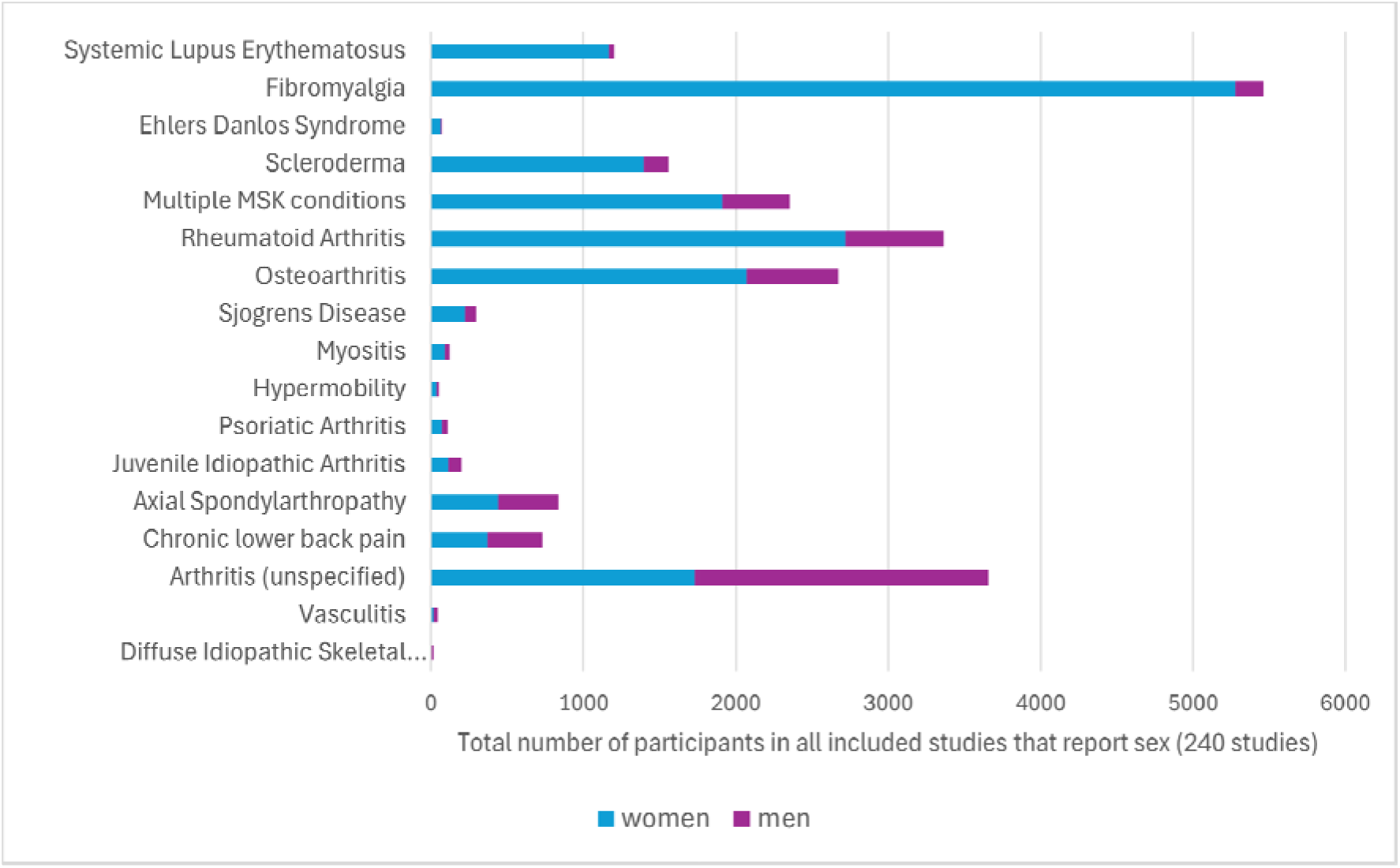

### Data charting

The data charting template was piloted by presenting the initial findings to a PPIE workshop group. In March 2024, a group of children and young people (n=7) reviewed this initial data and shared their opinions on which elements mattered to them and what was missing. This highlighted several elements of interventions which participants felt were important to ensuring safety but were not being captured.

Amendments to the data charting template were made to address this, recognising the importance of physical safety, in the sense that those involved in delivery were appropriately qualified and experienced in MSK health, and psychological safety in feeling accepted and supported. The additional items included in the data extraction template included: i) the knowledge and experience of the facilitator specific to the intervention, ii) the level of supervision of the intervention, iii) the physical and social environment where the intervention was delivered (Appendix 3).

Data from included papers were extracted by one reviewer and checked by another (KF and GY). Disagreements were resolved by consensus, with an additional reviewer (ED). The extracted data were analysed using narrative methods due to the heterogeneity of the included studies (28).

## RESULTS

### Review objective1: Explore the evidence for existing interventions for MSK fatigue

#### Intervention types and participant numbers

The types of intervention explored were categorised as educational, nutritional, passive therapy, physical activity, psychological, or multi component. A description of individual interventions can be found in the Supplementary Material (Appendix 4).

- Educational: interventions that increased knowledge and understanding of a health condition, specific symptoms or self-management skills
- Nutrition: interventions that included dietary changes or supplements taken orally
- Physical activity: interventions based on intentional movement or exercise
- Passive therapy: interventions that were provided by a device or a practitioner and did not require physical or mental exertion by the participants e.g. massage, acupuncture
- Psychological: interventions that addressed thoughts, feelings and patterns of behaviour
- Multi-component: including two or more non-pharmacological interventions

In general, the studies which focused primarily on fatigue had smaller numbers of participants. Across 268 included studies there were a total of 24,621 participants, 5,571 of whom participated in studies where fatigue was the primary focus. In studies where fatigue was of secondary interest, the most common primary focus was pain.

Most studies comprised a single category of intervention, however 76 studies reported on the use of multi-component interventions. The most common elements of multi-component interventions were physical activity, education and psychological support.

**Table 2.** Intervention types and participant numbers from included studies.

|  | Studies with a primary focus on fatigue<br>(*participants*) |  | All included studies<br>(*participants*) |  |
| --- | --- | --- | --- | --- |
|  | Single component | Multi-component | Single component | Multi-component |
| Physical activity | 26 (2191) | 13 (1489) | 91 (5915) | 60 (6836) |
| Passive therapy | 13 (733) | 1 (23) | 44 (5164) | 18 (1878) |
| Psychological | 9 (784) | 6 (917) | 31 (2971) | 40 (5059) |
| Educational | 6 (313) | 10 (1030) | 15 (1718) | 56 (5768) |
| Nutritional | 2 (45) | 1 (55) | 11 (347) | 5 (354) |
| **Total** | **56 (4066)** | **16 (1705)** | **192 (16115)** | **76 (8506)** |

The number of participants varied significantly with multicomponent studies tending to be larger than those reporting on a single component intervention. Physical activity interventions were the most studied when fatigue reduction was either the primary or secondary focus and regardless of whether a single or multi-component intervention was being reported. Both passive therapeutical interventions and psychological interventions were included in many studies of single interventions, however, passive therapeutic interventions appeared less frequently as part of a multi-component study. Educational interventions were the basis of only a small number of single component studies but appeared frequently in multi-component studies. Nutritional interventions were reported in fewer studies than any other type of intervention and had fewer participants.

#### Changes over time in the evidence being collected

There has been a notable increase in the number of studies that measure fatigue since 2007, likely related to the inclusion of fatigue in the core outcome set following the 8^th^ OMERACT meeting (29). Fatigue as a primary focus of studies has also increased but at a slower rate. There has been a slight increase in multicomponent interventions, but few focus primarily on fatigue and investigations of single component interventions remain the most common.

#### Geographical location of studies

The 268 included studies originated from 42 different countries across six continents. Most studies had a European origin (n=153) and the countries associated with the greatest number of eligible studies were the United States of America (n=49), Turkey (n=42) and Spain (n=24).

### Review objective 2: Explore the characteristics of included participants and identify who is missing

#### Sex

The sex of participants was reported in 240 of the included studies, of which 63 focused primarily on fatigue. Approximately 28% of research participants were men; however, the spread of participants by sex was uneven and skewed when viewed either by the intervention type or condition. Men were in the minority in all intervention types, most notably in nutrition, education and psychology.

Eighty-six studies only recruited women, no studies recruited exclusively men and 16 of the 240 studies had a majority of men taking part. Men have approximately equal representation in the research data relating to axial spondylarthritis (AxSpA), chronic lower back pain (CLBP), vasculitis and diffuse idiopathic skeletal hyperostosis (DISH). There are also similar numbers of men and women in research related to unspecified forms of arthritis, although this data came mainly from a small number of studies which explored the MSK health of veterans.

In studies relating to fibromyalgia, systemic lupus erythematosus (SLE), Ehlers Danlos Syndrome (EDS) and scleroderma, only a very small number of men were included, with many studies recruiting only women and some excluding men from participating. The uneven representation of men and women in the included studies may be due in part to sex imbalances in the distribution of individual conditions, but this alone does not explain the disparity.

#### Life stage of participants

The age of participants was reported in 263 of the included studies. Most studies included only adult participants (those aged 18 years or over), with 26 stratifying their results so findings relating to older adults (those aged over 65 years) could be identified, and a further four including exclusively older adults.

Children and young people were defined as those younger than 25 years of age to allow for variation in the timing of transition from paediatric to adult rheumatology services and to acknowledge the challenges associated with this transition (30,31). Two studies included both adult and child participants and a further 12 reported only on children and young people. In all age groups, physical activity was the most explored intervention. It was notable that there were no educational or multi-component interventions designed specifically for children and young people.

#### Clinical and demographic data

Clinical and demographic data of participants was collected in 243 of the included studies (Figure 3). The data collected most frequently was the socioeconomic status of study participants (n=129). Several studies discussed the impact of wider determinants on health literacy, patient activation and access to resources. Most participants were from medium or higher income households and had a higher than average level of formal education.

**Figure 3.**
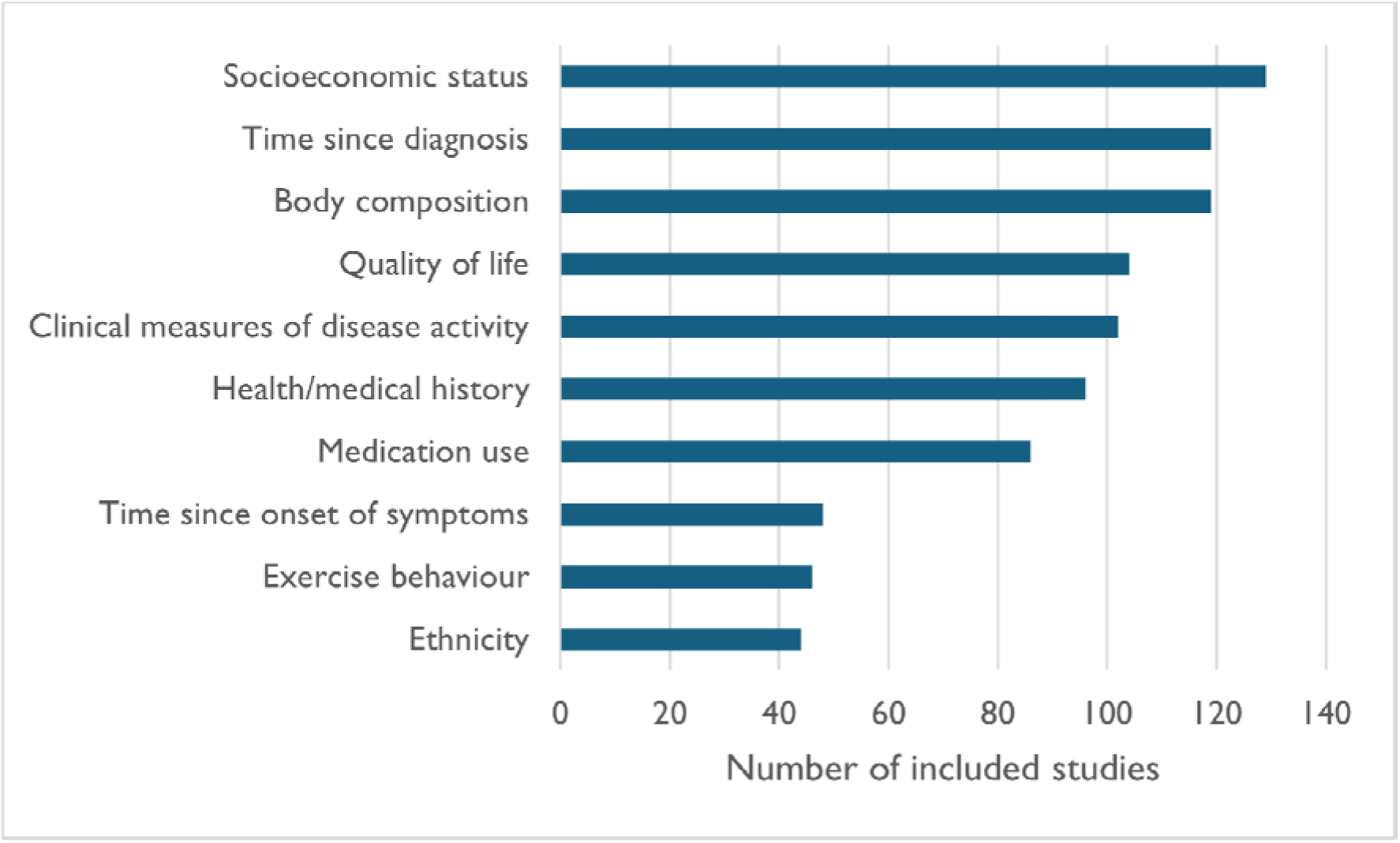

**Figure 4.**
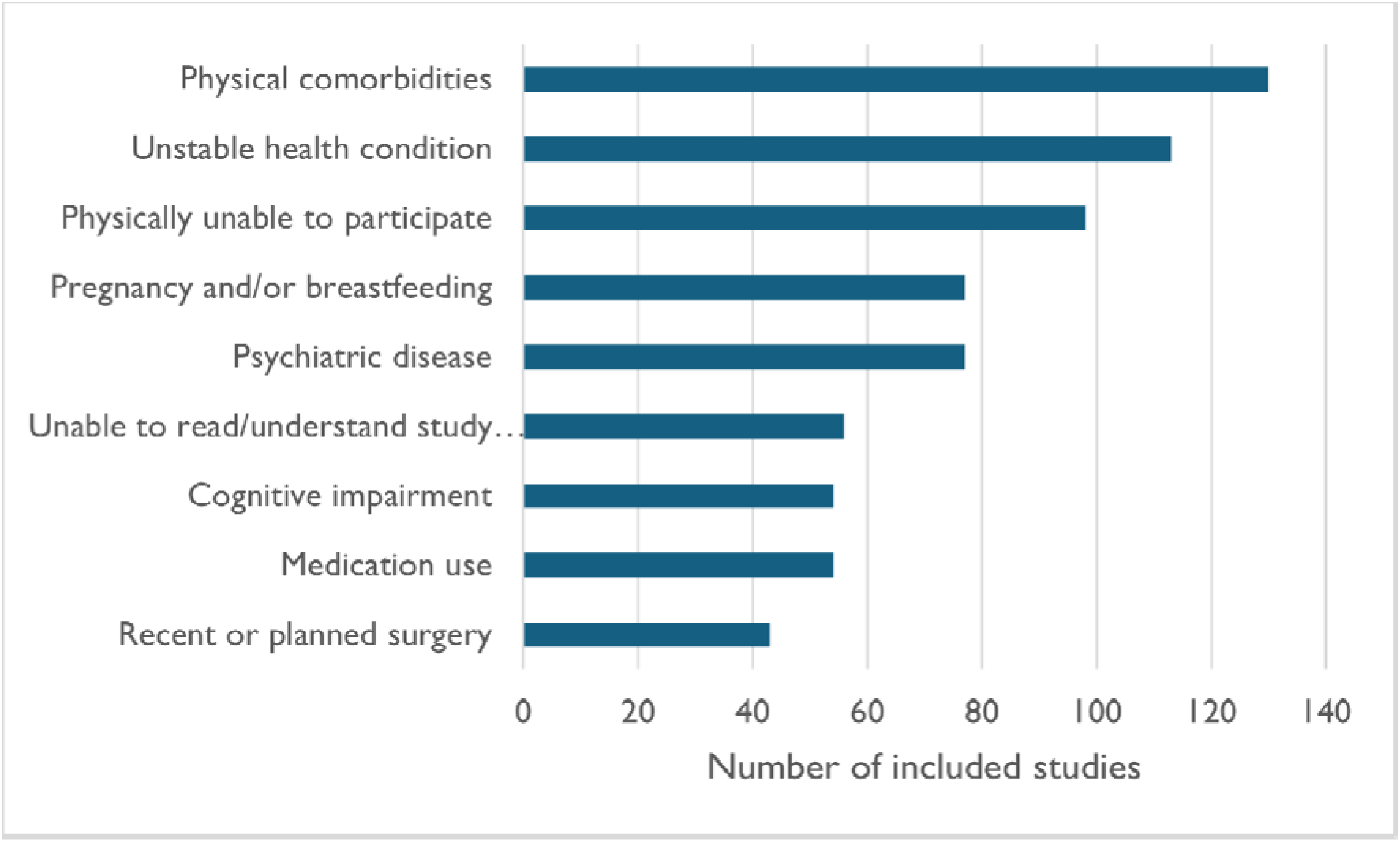

Data on body composition (height, weight, body mass index) was collected in almost half (n=119) of the included studies. In most cases the reasoning for this was not explained in the study, but a minority discussed the role of overweight and obesity in contributing to inflammation, potentially worsening fatigue.

Data on the time since diagnosis was also collected in 119 studies, which was noticeably distinct from time since onset of symptoms. Those studies which collected data on both often stated the impact of delayed diagnosis on the physical and mental wellbeing of patients.

It is significant that ethnicity data was collected in a relatively small number of studies (n=44), as there is evidence that some ethnic groups are underrepresented in research and also that cultural sensitivity is needed to ensure that interventions are acceptable to the target population (32). In the studies that did collect data about ethnicity, most participants were white Europeans.

#### Exclusion criteria

The criteria for excluding people from participation was reported by 222 included studies. This was explored to understand who is not represented in the current evidence base. The most common exclusion criteria were physical comorbidities or unstable health conditions reported in more than half of the included studies. Pregnancy and breastfeeding were also common reasons for excluding people, which is significant because most participants were women and many of the frequently studied conditions such as SLE, RA and fibromyalgia are prevalent in women of childbearing age.

Many studies excluded people who were living with cognitive impairment or unable to read/understand the study information. There are many reasons why a person may not be able to understand given information such as spoken languages, health literacy or disability, however the nature of this was rarely explored.

Other exclusion criteria included a history of substance misuse (n=11) or Body Mass Index (BMI) either above (n=7) or below (n=2) a stated value.

### Review objective 3: Explore the theoretical basis for existing interventions

The theoretical basis included the mechanism(s) through which the study authors theorised that the intervention was expected to reduce fatigue. This was stated in 68 of the included studies, including 27 studies where the aim of the intervention was primarily to reduce fatigue (Appendix 5). The stated mechanisms were categorised as being biochemical, physical and/or psychological in nature. The majority (n=56) explored a single mechanism; however, some studies considered two distinct mechanisms of action for reducing fatigue arising from their intervention.

**Table 3.** The frequency of mechanisms in relation to the category of intervention.

| **Mechanism category** | **Included studies** | **Intervention category** |  |  |  |  |
| --- | --- | --- | --- | --- | --- | --- |
|  |  | **Educational** | **Nutritional** | **Passive therapy** | **Physical activity** | **Psychological** |
| **Biochemical** | 30 | 2 | 5 | 15 | 13 | 2 |
| **Physical** | 28 | 1 | 2 | 8 | 18 | 3 |
| **Psychological** | 22 | 9 | 1 | 1 | 12 | 11 |

Biochemical mechanisms were described in 30 studies, most often in condition-specific interventions, in particular fibromyalgia (n=17). Potential mechanisms discussed included stimulation of the endocrine system by increasing neurotransmitter activity, regulating cortisol levels, reducing oxidative stress, and regulating metabolic activity. Other studies considered improvement of microcirculation, sympathetic nervous system regulation, immune system support and reduction of inflammation, specifically reducing obesity-induced inflammation, as potential biochemical mechanisms.

Physical mechanisms for reducing fatigue were most frequently explored in physical activity interventions and were discussed in 28 studies. Potential mechanisms included muscle relaxation, pain reduction, improved sleep quality, reduced BMI, increased aerobic capacity and increased muscle strength. Many conditions were represented in these studies, including CLBP, RA, AxSpA, SLE, psoriatic arthritis (PsA), juvenile idiopathic arthritis (JIA), fibromyalgia, scleroderma, Sjögren’s Disease, myositis and hypermobility.

Psychological mechanisms for reducing fatigue were explored in 22 studies across a range of MSK conditions. Many studies included participants with different conditions (n=7) and the most frequently studied specific condition was RA. The proposed mechanisms of action included general positive health management behaviours and specific actions such as increased knowledge, peer support, validation, increased self-efficacy, reduction of stress, acceptance and fear reduction.

### Review objective 4: Explore the design and outcome measures used by the included studies

In total, 268 records of unique studies were included in this review, of which 72 primarily focused on fatigue. The predominant methodology was quantitative (n=254) and the remaining studies used mixed methodology (n=14). Two of the quantitative studies were published alongside one or more complementary qualitative papers. These associated papers were grouped as single studies and data extracted from them accordingly, to avoid duplication while ensuring that rich contextual data were captured. The majority (9/14) of studies which used mixed methodology to explore the impact of their intervention were primarily focused on fatigue.

#### Fatigue measurement tools

A total of 35 tools were used to measure fatigue in the included studies, with 24 measurement tools used to assess those interventions specifically intended to reduce MSK fatigue (Appendix 6). Fifteen tools were identified that were specific to a single condition and 20 were validated for use in a range of health conditions. An age-specific tool, the Paediatric Quality of Life (PedQoL) was used to measure fatigue in 8 of the 12 studies that included children and young people.

### Review objective 5: Explore the location of existing interventions and the training and skills of those who delivered them

#### Location of delivery

The location where interventions were delivered was reported in 186 of the included studies. The most frequently reported locations were hospital outpatient clinics (n=80) and participant homes (n=59), with a minority (n=27) being delivered across multiple locations. Interventions were delivered both virtually and in person, with online delivery, telephone calls or text messages being used in 36 studies. Outside of hospitals, interventions were delivered in healthcare settings such as GP practices, clinics and university departments (n=23), and in community settings including gyms, swimming pools, community buildings and outdoor spaces (n=25). Two studies reported on interventions delivered in the participant’s workplace.

#### Intervention facilitators

Interventions were delivered by healthcare professionals, complementary medicine practitioners, physical activity instructors, researchers and peer support networks. The profession or role of the intervention facilitators was reported in 190 of the included studies and the majority of these (n=144) were delivered by one or more healthcare professionals.

In addition to details of the profession or experience of the intervention facilitators, 43 studies provided further details on the specific training which those delivering the intervention completed to support their role. Most interventions were delivered in a group format, although a notable minority (n=30) were self-guided (see Figure 5).

**Figure 5.**
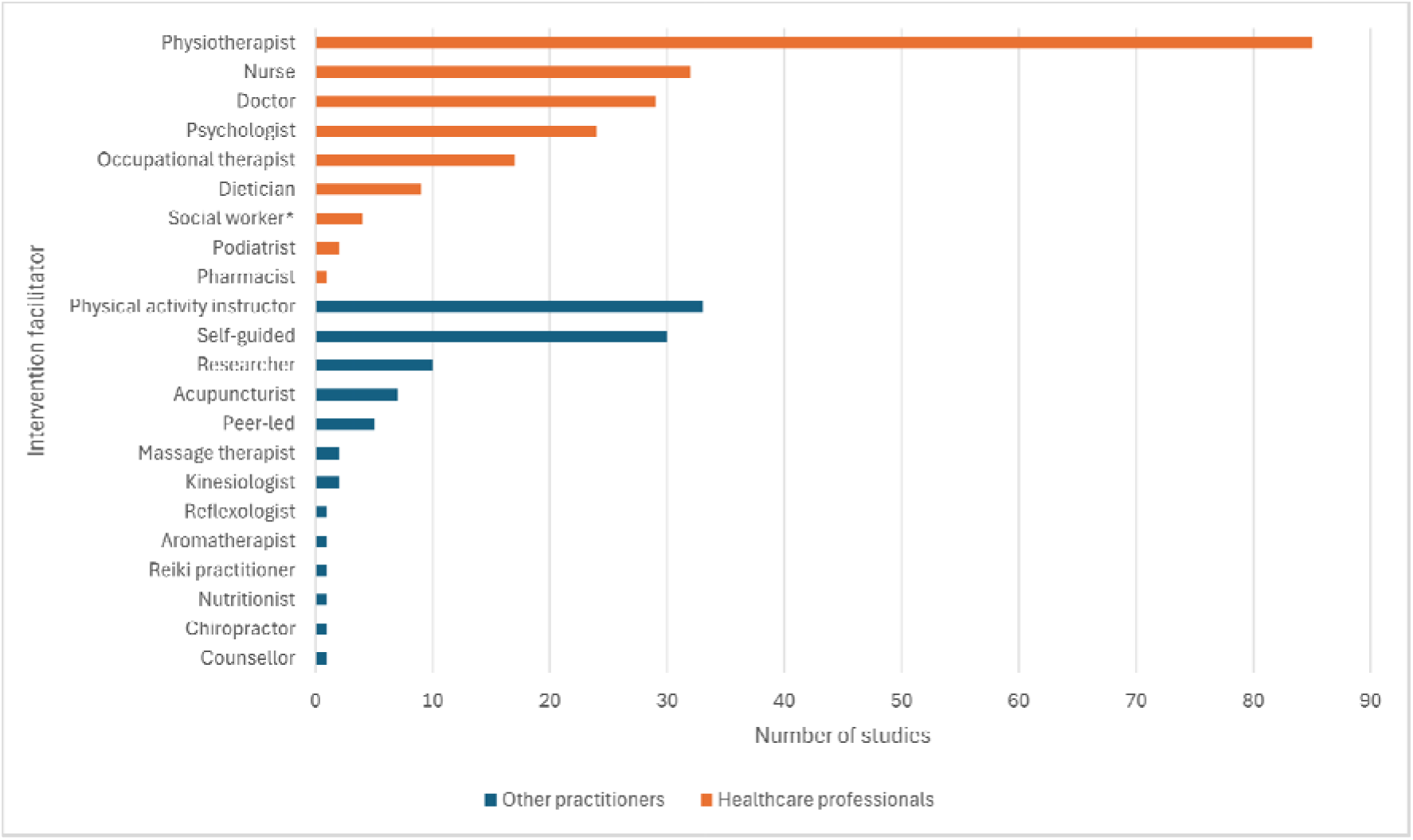

## DISCUSSION

Physical activity is the most studied intervention for fatigue, on its own and as part of multi-component interventions. The efficacy and safety of physical activity for fatigue management are well-evidenced (33), however, people with MSK conditions are less likely to be physically active than the general population (34,35). A PPIE workshop of children and young people explored the initial findings of this review, highlighting safety as the most important factor when determining the acceptability of a physical activity intervention. This included trust in the facilitator and adequate supervision underpinned by developmentally appropriate skills to feel physically safe, as well as environments that were appropriate to their age and condition to feel psychologically safe. This focus on trust and safety have been previously reported in qualitative research with adults (36). Further to this, adult PPIE contributors raised practical challenges around participation, in particular the importance of being offered interventions later in the day as fatigue was often worse during mornings.

MSK conditions explored in the included studies were largely representative of the prevalence of these conditions in the general population (37). Fibromyalgia was notable as the most frequently studied specific condition, and the condition where the mechanism of the intervention was most often described. This may reflect the prioritisation of fatigue as a symptom in this population or a greater focus on non-pharmacological management as recommended in EULAR guidelines (38,39).

Data were regularly collected on the socioeconomic status and body composition of participants, likely due to recognition that these factors represent disparities in MSK health. By contrast, ethnicity data were gathered infrequently. Where ethnicity of participants was reported, the majority were White Europeans, which is significant because research in England has found that people with Pakistani or Black Caribbean heritage are disproportionately affected by MSK conditions (40) and a further study found that people from some ethnic backgrounds were less likely to achieve disease remission within 3 months of treatment (41) A greater understanding of the wider determinants of MSK health is important to ensure that interventions are being developed which meet the needs of those most impacted by MSK fatigue.

The location where interventions were delivered was included in the data extraction at the suggestion of the PPIE contributors, as location was deemed important in relation to accessibility, physical and psychological safety. Most studied interventions were delivered by one or more healthcare professionals in clinical settings. Although this review did not collect cost data, it should be considered that the cost-effectiveness of different types of intervention may not be equal (42), which may impact overall feasibility and sustainability. There is currently a lack of evidence addressing the cost-effectiveness of non-pharmacological interventions which needs to be addressed in order to determine the optimal framework for fatigue management. (33).

A further aim of this scoping review was to highlight gaps in the evidence base to better understand who is not represented by the existing research and areas for potential interventions that were unexplored. There appears to be a general understanding of fatigue as multidimensional, with contributing factors varying between individuals and changing over time (43), however, only a minority (n=76) of included studies approached the management of fatigue holistically by investigating multicomponent interventions.

Activity pacing is a goal-directed behavioural process of supporting people to make decisions and plan how and where to distribute their available energy across daily activities (44) and was widely cited by PPIE contributors as a technique that had been recommended to them. It is the focus of only four of the included studies, though several others with educational components included pacing as one of many components. This relative paucity of evidence is surprising given the prevalence of this technique in clinical discussions, however, a recent meta-analysis of the effects of activity pacing in conditions associated with fatigue (45) found only six eligible papers, of which only two included participants with an MSK condition (46,47).

The role of nutrition in fatigue management was of particular interest to PPIE contributors from all age groups, however, only 16 studies included a nutritional element, and it was the least studied intervention type across all conditions and age groups. PPIE contributors discussed a lack of guidance on nutrition in relation to their condition and most had undertaken personal research to find recommendations through non-clinical avenues such as peer support groups and social media channels. This interest in nutrition may be because it is seen as an accessible resource and within the control of the patient. There are many challenges associated with designing high quality nutritional studies, such as accurate measurement of dietary intake and the influence of confounding factors (48), which may explain why evidence in this area is limited. Further research and comprehensive guidelines in this area would likely be welcomed by people with chronic MSK conditions.

The limited data specific to children or older adults is significant because of the different priorities and needs at different life stages. This was particularly highlighted by the children and young people in the PPIE workshops who felt that the complex relationships between education, health and social life needed to be considered when offering potential interventions.

Exclusion criteria are important to ensuring patient safety and data reliability in trials. Physical comorbidities were commonly cited as reasons for exclusion, meaning that those with more complex physical health needs were underrepresented. Psychiatric disease, cognitive deficit and pregnancy/breastfeeding were also common exclusion criteria in the included studies. Non-pharmacological interventions are likely to be of benefit to these populations, so it is important to ensure that they have opportunities to participate in research and that their needs are being considered when designing interventions for implementation. A further challenge will be to address the evidence to practice gap by considering innovative ways to overcome common barriers to implementation.

PPIE contributors were engaged at multiple stages during the review process to ensure that the review questions were relevant. Being responsive to the contributions of PPIE groups has ensured that the findings presented represent the priorities of patients and clinicians. Additionally, it has deepened the research team’s understanding of the nuanced experiences of fatigue and the specific challenges that individuals face at different stages of their lives.

Key themes which emerged from the discussions included the need for research findings to be easily accessible to patients from reliable sources and wanting to be offered or signposted to a range of options for fatigue management as part of standard rheumatology care. The lived experience of the PPIE groups added valuable insight into the nature and challenges of MSK-fatigue and their input had a significant impact on the search strategy, data extraction process and prioritisation of review findings.

The scoping review methodology captured rich data from a broad range of study types which would not be included in a traditional systematic review. This methodology is more appropriate for mapping the existing evidence base as the wide inclusion criteria reveals where evidence overlaps and identifies gaps (49). However, the number of studies included meant that extensive categorisation was necessary to synthesise a large, heterogeneous data set. This can lead to a loss of nuance in areas such as the description of the interventions, for example an exercise intervention may be psychologically informed and therefore more multidimensional than it otherwise appears.

A further limitation was the exclusion of studies that were not published in English language. This potentially limited alternative views on non-pharmacological management and the diversity of the participants (50). The included studies mainly originated from Europe and North America, meaning that traditional western views of medicine and healthcare dominate findings. Including other languages and databases would potentially reveal a greater depth of evidence for some of the areas which appear as underrepresented.

## CONCLUSIONS

There are a huge number of non-pharmacological interventions that have been studied to explore their impact on fatigue. Despite variation in the mechanisms of fatigue and the interventions across the range of MSK conditions, there is strong agreement that both the causes and the experience of fatigue are multidimensional. Physical activity interventions to improve muscle strength and aerobic capacity were the mostly widely studied but more research is needed to understand how these interventions can be made accessible and acceptable for those groups who are living with highly intrusive symptoms of fatigue.

The involvement of the PPIE groups in exploring the review findings has demonstrated a potential disconnect between the priorities of patients and those of researchers in terms of both preference for intervention types and information sharing needs.

Fatigue is frequently considered in studies of symptom management for MSK conditions, and the causes and experience of fatigue are widely acknowledged as multidimensional. However, there are relatively few studies which explore how approaches can be combined to achieve person-centred fatigue management.

Gaps in the evidence base have been identified, including limited evidence on appropriate interventions for managing fatigue in children and young people. While the mechanism of action of some interventions may be similar across the lifespan, the criteria which determines whether an intervention is both accessible and acceptable is likely to change. Future studies should also explore participants’ perceptions in relation to their physical and psychological safety.

Underrepresented groups included men, people with complex or unstable health conditions and those living with cognitive or intellectual impairment. To effectively tailor interventions to the needs of individuals it is important to understand how these characteristics may interact with health needs.

## Data Availability

All data produced in the present study are available upon reasonable request to the corresponding author

## AUTHOR CONTRIBUTIONS

Project funding application by ED and PM with support from FC, JP, GM, CC, BF and ES. Review conducted by KF and GY with contributions by ED, FC and JP. Article drafted by KF in discussion with GY and ED, structure and content reviewed by CC, FC, EE, BF, GM, PM, JP and ES.

## ACKNOWLEDGEMENTS

The development of this scoping review search strategy was supported by Specialist Subject Librarian Pauline Shaw from the University of the West of England library. The review team is also incredibly grateful for the contributions of the stakeholders who attended the Patient and Public Involvement and Engagement workshops and to the patient organisation YourRheum for facilitating PPIE workshops with children and young people living with arthritis.

