## Appendix 1 for "A scoping review of non-pharmacological interventions for managing fatigue across the lifespan of people living with chronic musculoskeletal conditions"

Appendix 1. GRIPP2 Short form reporting checklist

| Section and topic | Item | Reported on page No |
| --- | --- | --- |
| 1: Aim | Report the aim of PPI in the study | 3 |
| 2: Methods | Provide a clear description of the methods used for PPI in the study | 3 |
| 3: Study results | Outcomes—Report the results of PPI in the study, including both positive and negative outcomes | 5, 12 |
| 4: Discussion and conclusions | Outcomes—Comment on the extent to which PPI influenced the study overall. Describe positive and negative effects | 13, 14 |
| 5: Reflections/critical perspective | Comment critically on the study, reflecting on the things that went well and those that did not, so others can learn from this experience | 13 |
