## Appendix 2 for "A scoping review of non-pharmacological interventions for managing fatigue across the lifespan of people living with chronic musculoskeletal conditions"

**Appendix 1: Search terms and search strategy overview**

|  | **Population** | **Exposure** | **Outcome** |
| --- | --- | --- | --- |
| **Search terms** | musculoskeletal condition  musculoskeletal disease  inflammatory arthritis  rheumat*  osteoarthritis  fibromyalgia  juvenile idiopathic arthritis  juvenile chronic arthritis  spondyloarth*  systemic sclerosis  Sjogren’s syndrome  lupus  psoriatic arthritis  spondylitis  scleroderma  *myositis  immune mediated necrotising myopathy  anti synthetase syndrome  vasculitis  giant cell arteritis  Takayasu’s arteritis  polyarteritis nodosa  granulomatosis with polyangiitis  microscopic angiitis  Behcet syndrome  Wegener syndrome  Churg-Strauss syndrome  low* back pain  Ehlers Danlos  hypermobility | non-pharmacological intervention  physical activity  exercis*  mindfulness  cognitive behavioural therapy  psycho-educational  psychosocial  education*  complementary  holistic therap*  nutrition  diet  massage  nurse-led  physiotherapy  occupational therapy  electro-physical modalities  thermotherapy  manual therapy  balneotherapy  tai chi  yoga  reflexology  aromatherapy  chiropractic  acupuncture  electroacupuncture  low level laser therapy  electric stimulation therapy  hyperthermia  walk*  pacing  hydrotherapy  activity diar*  swim*  dry needl* | fatigue |
| **Databases** | - EBSCO (AMED, CINAHLPlus, MEDLINE, PsycINFO) - Ovid (Embase) - Scopus - Cochrane Database | | |
| **Inclusion criteria** | - Primary research study - Published in a peer reviewed journal - Available in English language - Participants have one or more chronic musculoskeletal conditions - Participants experience fatigue at baseline - Published during or after 2007 - Describes an intervention to manage MSK condition symptoms, with fatigue reduction as a primary or secondary outcome | | |
| **Exclusion criteria** | - Reviews, protocols, opinion pieces, editorials, case reports, case series, observational cohort studies - Pharmacological interventions - No intervention is described - Muscle fatigue rather than global fatigue is examined - No data is available on factors associated with intervention success (theoretical mechanism of intervention OR characteristics of participants OR characteristics of clinicians delivering interventions) | | |

**Search strategy**

**EBSCO (AMED, CINAHLPlus, MEDLINE, PsycINFO)**

S1	TITLE (“musculoskeletal condition” OR “musculoskeletal disease” OR “inflammatory arthritis” OR rheumat* OR osteoarthritis OR fibromyalgia OR “juvenile idiopathic arthritis” OR “juvenile chronic arthritis” OR spondyloarth* OR “systemic sclerosis” OR “Sjogren’s syndrome” OR lupus OR “psoriatic arthritis” OR spondylitis OR scleroderma OR *myositis OR “immune-mediated necrotising myopathy” OR “anti-synthetase syndrome” OR vasculitis OR “giant cell arteritis” OR “Takayasu’s arteritis” OR “polyarteritis nodosa” OR “granulomatosis with polyangiitis” OR “microscopic angiitis” OR “Behcet syndrome” OR “Wegener syndrome” OR “Churg-Strauss syndrome” OR “low* back pain” OR “Ehlers Danlos” OR hypermobility) OR ABSTRACT (“musculoskeletal condition” OR “musculoskeletal disease” OR “inflammatory arthritis” OR rheumat* OR osteoarthritis OR fibromyalgia OR “juvenile idiopathic arthritis” OR “juvenile chronic arthritis” OR spondyloarth* OR “systemic sclerosis” OR “Sjogren’s syndrome” OR lupus OR “psoriatic arthritis” OR spondylitis OR scleroderma OR *myositis OR “immune-mediated necrotising myopathy” OR “anti-synthetase syndrome” OR vasculitis OR “giant cell arteritis” OR “Takayasu’s arteritis” OR “polyarteritis nodosa” OR “granulomatosis with polyangiitis” OR “microscopic angiitis” OR “Behcet syndrome” OR “Wegener syndrome” OR “Churg-Strauss syndrome” OR “low* back pain” OR “Ehlers Danlos” OR hypermobility)

S2	TITLE (“non-pharmacological intervention” OR “physical activity” OR exercis* OR mindfulness OR “cognitive behavioural therapy” OR “psycho-educational” OR psychosocial OR education* OR complementary OR “holistic therap*” OR nutrition OR diet OR massage OR “nurse-led” OR physiotherapy OR “occupational therapy” OR “sports rehabilitation therapy” OR “electro-physical modalities” OR thermotherapy OR “manual therapy” OR balneotherapy OR “tai chi” OR yoga OR reflexology OR aromatherapy OR chiropractic OR acupuncture OR electroacupuncture OR “low level laser therapy” OR “electric stimulation therapy” OR hyperthermia OR walk* OR pacing OR hydrotherapy OR “activity diar*” OR swim* OR “dry needl*”) OR ABSTRACT (“non-pharmacological intervention” OR “physical activity” OR exercis* OR mindfulness OR “cognitive behavioural therapy” OR “psycho-educational” OR psychosocial OR education* OR complementary OR “holistic therap*” OR nutrition OR diet OR massage OR “nurse-led” OR physiotherapy OR “occupational therapy” OR “sports rehabilitation therapy” OR “electro-physical modalities” OR thermotherapy OR “manual therapy” OR balneotherapy OR “tai chi” OR yoga OR reflexology OR aromatherapy OR chiropractic OR acupuncture OR electroacupuncture OR “low level laser therapy” OR “electric stimulation therapy” OR hyperthermia OR walk* OR pacing OR hydrotherapy OR “activity diar*” OR swim* OR “dry needl*”)

S3	TITLE (fatigue) OR ABSTRACT (fatigue)

S4	S1 AND S2 AND S3

S5	Limit S5 to;

- Publication date during or after 2007
- English language
- Peer reviewed

**3.2 Ovid (EMBASE)**

#1	(musculoskeletal condition OR musculoskeletal disease OR inflammatory arthritis OR rheumat* OR osteoarthritis OR fibromyalgia OR juvenile idiopathic arthritis OR spondyloarth* OR systemic sclerosis OR Sjogrens syndrome OR lupus OR psoriatic arthritis OR spondylitis OR scleroderma OR myositis OR immune-mediated necrotising myopathy OR anti-synthetase syndrome OR vasculitis OR giant cell arteritis OR Takayasus arteritis OR polyarteritis nodosa OR granulomatosis with polyangiitis OR microscopic angiitis OR Behcet syndrome OR Wegener syndrome OR Churg-Strauss syndrome OR low* back pain OR Ehlers Danlos OR hypermobility).ti OR (musculoskeletal condition OR musculoskeletal disease OR inflammatory arthritis OR rheumat* OR osteoarthritis OR fibromyalgia OR juvenile idiopathic arthritis OR spondyloarth* OR systemic sclerosis OR Sjogrens syndrome OR lupus OR psoriatic arthritis OR spondylitis OR scleroderma OR myositis OR immune-mediated necrotising myopathy OR anti-synthetase syndrome OR vasculitis OR giant cell arteritis OR Takayasus arteritis OR polyarteritis nodosa OR granulomatosis with polyangiitis OR microscopic angiitis OR Behcet syndrome OR Wegener syndrome OR Churg-Strauss syndrome OR low* back pain OR Ehlers Danlos OR hypermobility).ab.

#2	Limit 1 to (English language and yr=”2007”)

#3	(fatigue).ti. or (fatigue).ab.

#4	Limit 2 to (English language and yr=”2007”)

#5	(non-pharmacological intervention OR physical activity or exercis* OR mindfulness OR cognitive behavioural therapy OR psycho-educational OR psychosocial OR education* OR complementary OR holistic therp* OR nutrition OR diet OR massage OR nurse-led OR physiotherapy OR occupational therapy OR sports rehabilitation therapy OR electro-physical modalities OR thermotherapy OR manual therapy OR balneotherapy OR tai chi OR yoga OR reflexology OR aromatherapy OR chiropractic OR acupuncture OR electroacupuncture OR low level laser therapy OR electric stimulation therapy OR hyperthermia OR walk* OR pacing OR hydrotherapy OR activity diar* OR swim* OR dry needl*).ti. OR (non-pharmacological intervention OR physical activity or exercis* OR mindfulness OR cognitive behavioural therapy OR psycho-educational OR psychosocial OR education* OR complementary OR holistic therp* OR nutrition OR diet OR massage OR nurse-led OR physiotherapy OR occupational therapy OR sports rehabilitation therapy OR electro-physical modalities OR thermotherapy OR manual therapy OR balneotherapy OR tai chi OR yoga OR reflexology OR aromatherapy OR chiropractic OR acupuncture OR electroacupuncture OR low level laser therapy OR electric stimulation therapy OR hyperthermia OR walk* OR pacing OR hydrotherapy OR activity diar* OR swim* OR dry needl*).ab.

#6	Limit 5 to (English language and yr=”2007”)

#7	2 and 4 and 6

**3.3 SCOPUS**

#1	(TITLE-ABS("non pharmacological intervention" OR "physical activity" OR exercis* OR mindfulness OR "cognitive behavioural therapy" OR psycho-educational OR psychosocial OR education* OR complementary OR “holistic therap*” OR nutrition OR diet OR massage OR "nurse-led” OR physiotherapy OR "occupational therapy" OR “sports rehabilitation therapy” OR "electro-physical modalities" OR thermotherapy OR "manual therapy" OR balneotherapy OR "tai chi" OR yoga OR reflexology OR aromatherapy OR chiropractic OR acupuncture OR electroacupuncture OR "low level laser therapy" OR "electric stimulation therapy" OR hyperthermia OR walk* OR pacing OR hydrotherapy OR “activity diar*” OR swim* OR “dry needl*”)

#2	Limit #1 to:

- Publication during or after 2007
- English language

#3	TITLE-ABS(fatigue)

#4	Limit #3 to:

- Publication during or after 2007
- English language

#5	TITLE-ABS(“musculoskeletal condition” OR “musculoskeletal disease” OR “inflammatory arthritis” OR rheumat* OR osteoarthritis OR fibromyalgia OR “juvenile idiopathic arthritis” OR spondyloarth* OR “systemic sclerosis” OR “Sjogrens syndrome” OR lupus OR “psoriatic arthritis” OR spondylitis OR scleroderma OR *myositis OR “immune-mediated necrotising myopathy” OR “anti-synthetase syndrome” OR vasculitis OR “giant cell arteritis” OR “Takayasus arteritis” OR “polyarteritis nodosa” OR “granulomatosis polyangiitis” OR “microscopic angiitis” OR “Behcet syndrome” OR “Wegener syndrome” OR “Churg-Strauss syndrome” OR “low* back pain” OR “Ehlers Danlos” OR hypermobility)

#6	Limit #5 to:

- Publication during or after 2007
- English language

#7	#2 AND #4 AND #6

**3.4 COCHRANE LIBRARY DATABASE**

ID	Search terms	Results

#1	("non pharmacological intervention" OR "physical activity" OR exercis* OR mindfulness OR "cognitive behavioural therapy" OR psycho-educational OR psychosocial OR education* OR complementary OR “holistic therap*” OR nutrition OR diet OR massage OR "nurse-led” OR physiotherapy OR "occupational therapy" OR “sports rehabilitation therapy” OR "electro-physical modalities" OR thermotherapy OR "manual therapy" OR balneotherapy OR "tai chi" OR yoga OR reflexology OR aromatherapy OR chiropractic OR acupuncture OR electroacupuncture OR "low level laser therapy" OR "electric stimulation therapy" OR hyperthermia OR walk* OR pacing OR hydrotherapy OR “activity diar*” OR swim* OR “dry needl*”):ti,ab

#2	(fatigue):ti,ab

#3	(“musculoskeletal condition” OR “musculoskeletal disease” OR “inflammatory arthritis” OR rheumat* OR osteoarthritis OR fibromyalgia OR “juvenile idiopathic arthritis” OR spondyloarth* OR “systemic sclerosis” OR “Sjogren’s syndrome” OR lupus OR “psoriatic arthritis” OR spondylitis OR scleroderma OR *myositis OR “immune-mediated necrotising myopathy” OR “anti-synthetase syndrome” OR vasculitis OR “giant cell arteritis” OR “Takayasu’s arteritis” OR “polyarteritis nodosa” OR “granulomatosis with polyangiitis” OR “microscopic angiitis” OR “Behcet syndrome” OR “Wegener syndrome” OR “Churg-Strauss syndrome” OR “low* back pain” OR “Ehlers Danlos” OR hypermobility):ti,ab

#4	#1 AND #2 AND #3

#5	Limit to:

- Publication date from Jan 2007 to Oct 2023
- English language
