## Appendix 3 for "A scoping review of non-pharmacological interventions for managing fatigue across the lifespan of people living with chronic musculoskeletal conditions"

**Appendix 3: Amended data extraction template**

|  | **Study number** | **1** | **2** | **3 etc.** |
| --- | --- | --- | --- | --- |
| **Review objective 1: Evidence for existing interventions** | Title |  |  |  |
|  | Lead author |  |  |  |
|  | Publication year |  |  |  |
|  | Country |  |  |  |
|  | Number of participants |  |  |  |
|  | Condition(s) included |  |  |  |
|  | Primary or secondary focus on fatigue |  |  |  |
|  | Category of intervention |  |  |  |
| **Review objective 2: Characteristics of participants** | Sex |  |  |  |
|  | Age |  |  |  |
|  | Clinical and demographic data collected |  |  |  |
|  | Exclusion criteria stated |  |  |  |
| **Review objective 3: Theoretical basis for interventions** | Description of intervention |  |  |  |
|  | Theoretical basis described to justify intervention |  |  |  |
| **Review objective 4: Design and outcome measures** | Fatigue measurement tool(s) used |  |  |  |
|  | Methodology |  |  |  |
| **Review objective 5: Location and facilitator training/skills** | Setting for intervention delivery |  |  |  |
|  | Profession/role of intervention facilitator(s) |  |  |  |
|  | Additional training given specific to studied intervention |  |  |  |
