## Appendix 4 for "A scoping review of non-pharmacological interventions for managing fatigue across the lifespan of people living with chronic musculoskeletal conditions"

**Appendix 4. Description of individual interventions**

| **Author** | **Year** | **Brief description of intervention** | **Intervention category** |
| --- | --- | --- | --- |
| Abou-Raya | 2014 | Participants randomised to; diet only, exercise only, diet + exercise or usual care groups | Nutritional; Physical activity |
| Aleksandrov | 2022 | Participants directed to do either <5-6000 steps/day (group 1) OR >7-8000 steps/day (group 2) | Physical activity |
| Alentorn-Geli | 2008 | 6-week aerobic exercise protocol and Whole body vibration while doing static positions on the machine with it switched on (WBV) (2/7), compared to 6-week aerobic exercise protocol (2/7) static postures on the WBV machine with it switched off , compared to usual care | Passive therapy |
| Alexanderson | 2014 | Progressive aerobic exercise and strength training 3/7 for 8 weeks | Physical activity |
| Al-Herz | 2008 | Physiotherapy guided strength exercises, Mobility exercises, stretching exercises | Physical activity |
| Anderson | 2007 | All patients were enrolled in a one year MDT outpatient treatment program combining pharmacologic and non pharmacologic treatment, including CBT coping skills classes, exercise, massage, nutritional counselling and electromedicine interventions. Eight coping skills classes inc. lectures, group discussion, group exercises, homework assignments and reading. One group had four additional psychotherapy sessions | Psychological |
| Aree-Ue | 2022 | 3-day health workshop. Intervention group were supported by volunteers within village setting, no additional support for control group | Psychological |
| Avaux | 2016 | Supervised training group received multidisciplinary information session about benefits of exercise in SLE, including practical information. Three hours exercise per week for 12 weeks, including endurance exercises (walking or cycling) and strengthening exercises (elastobands or weights), performed under the supervision of the multidisciplinary team.  Home training group also received multidisciplinary information session about benefits of exercise in SLE, including practical information. Three hours exercise per week for 12 weeks, including endurance exercises (walking or cycling) and strengthening exercises (elastobands or weights), performed at homeControl group did not participate in information session and were not asked to change level of physical activity | Educational; Physical activity |
| Azeez | 2020 | 3-month personalized exercise programme, prescribed by the study physiotherapist (CC) and had three sessions with the physio-therapist during the period of study. cardiovascular exercise prescribed (walking,cycling or swimming). Strength training for all major muscle groups | Physical activity |
| Bachmair | 2022 | PEP group - core components were formation of a problem statement, completion of activity diaries, goal setting, homework activities, review and feedback.  CBA group - completion of activity diaries, goal setting, homework activities, review and feedback.  Control group - Versus Arthritis booklet on fatigue. | Physical activity; Psychological |
| Baelz | 2023 | Chinese acupuncture for 30 minutes once a week for four consecutive weeks vs. sham acupuncture 30 minutes once a week for four consecutive weeks | Passive therapy |
| Bagdatli | 2015 | 6hr patient education- info about FM, exercise prescription, psychological education vs. 6hrs as per control plus 10 heated pool baths, and 10 mud pack applications | Passive therapy |
| Baglan Yentur | 2020 | Pilates: three 45min sessions, weekly for 8 weeks. Exercise balls and resistance elastic bands were used in the latter weeks of the treatment according to the patient’s condition. Breathing control, postural exercises, sitting exercises, and proprioceptive exercises were taught and applied to the patients. Stretching, relaxation, and postural exercises were used for cool-down.  Aerobic: three sessions per week for 8 weeks walking on treadmill. Maximum heart rate was calculated and followed with a monitor during the training. The aim was to reach 60-80% of MHR as training workload. The training began with under the workload for 5 min for warm-up, then continued with training workload for 20 min, and finished with cool-down for 5 min.  Combined: both protocols were used. Participants completed the aerobic exercises first, rested for 15 minutes and then completed the Pilates exercises. Three sessions per week for 8 weeks. | Physical activity |
| Baldi | 2022 | Consumption of Khorsan wheat products vs control wheat products | Nutritional |
| Bestas | 2022 | Group 1 - 20 sessions held 5 days a week over 4 weeks. Water-based Exercise- 60 minutes exercise 33c  Group 2 - 20 sessions held 5 days a week over 4 weeks. Land based exercise - 60 minutes inc 20 minutes therapeutic local massage of paravertebral muscles  Control group - 20 sessions held 5 days a week over 4 weeks. Balneotherapy- 20 minutes whole body spa 39c | Passive therapy; Physical activity |
| Bettoni | 2013 | 15 sessions of whole body cryotherapy (WBC) within a period of 3 weeks. Thirty seconds pre conditioning at -60C followed by 3 minutes exposure to -140C then 30 mins compulsory aerobic exercise (cycloergometer or treadmill) | Passive therapy |
| Bilberg | 2023 | Intervention group - 12 week exercise program including supervised cardiorespiratory high intensity interval exercise and strength exercise twice per week, plus an additional non supervised session of the patients choice  Control group - Information for physical activity according to general health recommendations and encouragement to be moderately active for at least 150 minutes per week | Physical activity |
| Bircan | 2008 | Group1 - Aerobic exercise three times per week for eight weeks. 20 minutes walking on treadmill, increasing to 30 minutes as tolerated by participant, 5 minutes of mild stretching before and after  Group 2 - Strength exercise three times per week for eight weeks. Supervised 30 minute progressive physical training programme in a group setting using free weights and body weights in a variety of positions, increasing weight and repetitions as tolerated by participant. 5 minutes warm up and 5 mins cool down | Physical activity |
| Bjersing | 2017 | 15 week exercise program performed twice a week. Exercise lasted for about 1 hour. Began with a 10 min warm-up period, then resistance exercise for legs, arms and hands and core stability, ended with stretching exercise. After 3-4 weeks load increased to 60% and then to 80% of 1RM. Explosive strength exercises for legs were included at 5 and 8 weeks. | Physical activity |
| Blaney | 2021 | Worry and Sadness program - 6 modules to be completed in order, approx 15 minutes each. Submission required at the end of each session in order to access the next | Psychological |
| Bogdanovic | 2015 | Group 1 - aerobic training on a bicycle ergometer for a period of 15 minutes, 3 times per week for 6 weeks  Group 1 - aerobic training on a bicycle ergometer for a period of 15 minutes, 3 times per week for 6 weeks | Physical activity |
| Bossen | 2013 | Join2move - baseline test, goal setting, time contingent physical activity and text messages to promote PA. Join2move intervention is a self-paced 9-week PA program in which a patients favourite recreational activity is gradually increased. Users select a central activity such as cycling, walking, or gardening; perform a 3-day self-test; and determine a short-term goal for the next 8 weeks. 8 tailored weekly modules are generated. | Physical activity |
| Braz | 2012 | Amitriptyline hydrochloride (25mg/d) vs. P. ginseng root extract (25mg/d) vs. placebo (talcum powder) | Nutritional |
| Britto | 2020 | Group 1 - Water based exercise  Water temperature was approx 33 C. Assisted stretching activity of the entire posterior and anterior muscle chain was per- formed with three repetitions and lasted 15 to 25 s for each muscle group. The strengthening exercises for the lower limbs were per- formed with a shin guard in three sets of 15 repetitions of each move- ment, and noodle floats were used for the upper limbs, also with three sets of 15 repetitions of each movement.  Group 2 - Land based exercise  Included warm-up, active stretching, strengthening, and relaxation under the supervision of a physical therapist | Physical activity |
| Broderick | 2014 | Four broad coping skills were taught across the ten 30- to 45-minute sessions: relaxation response, attention diversion techniques, altering activity and rest patterns as a way of increasing activity level, and reducing negative pain-related thoughts and emotions. Then self-guided practice at home with homework | Psychological |
| Broderick | 2016 | Pain Coping Skills training (CBT) - 10 individual weekly sessions delivered at the patient's GP surgery. Up to 4 could be delivered by telephone. The sessions were outlined in detail in a treatment manual and followed a format of review of home practice assigned at the previous session, instruction in a new coping skill, guided practice in that skill, and a home practice assignment. Homework assignments followed by review and problem-solving in the subsequent session. | Psychological |
| Callahan | 2008 | PACE (people with arthritis can exercise) program. 8 week program, 2x weekly for 1hour, intended to promote self-management of arthritis through exercise | Physical activity |
| Callahan | 2011 | Walk With Ease (WWE) - 6 week community based walking programme, WWE program content focuses on having all participants engage in motivational strategies, including goal setting, setting up action plans, monitoring progress, identifying rewards, and using social supports. Participants selected their own allocation.  Self-directed participants followed the program alone with a workbook as a guide.  Group participants lead by an instructor and, meeting 3 times per week for 1 hour. | Educational; Physical activity |
| Cano-Garcia | 2014 | Face to face group delivery of standardised educational intervention, including oral presentations and group work | Educational; Psychological |
| Carandang | 2022 | FAME (Fatigue and Activity Management Education)-ISS Six 90 minute meetings held weekly for 6 weeks. Electronic and paper copies of handbook provided | Educational |
| Carbonario | 2013 | After initial assessment, all participants were oriented about fibromyalgia, including symptoms and the importance  of physical exercises. Instructed in how to properly carry weights and about adequate postures. They also received a basic program consisting of 11 types of stretching exercises. They received a syllabus summarising the information and illustrating the exercises. Aerobic exercises were conducted on an ergometer bicycle at the participants' own speed for 30 minutes.  Intervention group received additional TENS (transcutaneous electrical nerve stimulation) - 150 Hz applied to bilateral tender points of trapezium and supraspinatus. | Passive therapy |
| Carbonell-Baeza | 2011 | Three sessions per week for 12  weeks. Two sessions per week were performed in a chest-high warm pool  for 45 minutes, and the third session included 45 minutes of activity in the exercise room and 90 minutes  of psychological-educational therapy.  The exercise sessions were supervised by a fitness specialist The psychological-educational sessions were conducted by a psychologist with experience in treating fibromyalgia patients. | Educational; Physical activity; Psychological |
| Carson | 2010 | Yoga of Awareness - 8 week programme, 120 minutes once per week. The protocol employed a  series of low intensity, low impact yoga poses which were modified for common pathophysiologic changes in FM. This was complemented by meditation and breathing exercises, yoga-based coping presentations, and group discussions | Physical activity |
| Carvalho | 2020 | The exergames group performed the program thrice perweek with each session lasting 1 hour. Six subgames of Wii Fit Plus were chosen for this group .  The control group performed the chain muscle stretching technique thrice per week with each session lasting 1 hour. This schedule was followed for the 7 weeks of the study. The positions were held during four deep and prolonged expirations. | Physical activity |
| Casanueva | 2014 | Weekly 1-h session of dry needling over the 18 tender points for a 6-week period. All dry needling procedures were performed by the same investigator, and the technique used (superficial dry needling or deep dry needling) was similar to the Baldry method, using sterile Ener-Qi acupuncture needles for the punction of the tender points of fibromyalgia | Passive therapy |
| Casanueva-Fernandez | 2012 | All participants continued current medical treatment and participated in four patient education sessions.  Intervention group received an additional weekly 1-h session for 8 weeks including: (1) Massage therapy for 15 min. (2) Ische- mic pressure upon the 18 tender points during a total time of about 25 min. (3) Aerobic exercise: using a stationary bicycle for 5 min and a treadmill for 5 min. (4) Thermal therapy using convective heat transfer with a 250-W infrared heat lamp for 10 min | Passive therapy; Physical activity |
| Cash | 2015 | The Mindfulness Based Stress Reduction (MBSR) group followed the format of the traditional program, meeting for weekly 2.5-hour sessions over 8 weeks with an experienced, trained MBSR instructor. Both formal and informal mindfulness practices were introduced including instruction/discussion, an attention-focusing technique, sitting meditation, and a series of simple yoga positions taught as a means of encouraging relaxed and focused movement. | Psychological |
| Castro Sanchez | 2019 | Dry needling group - 4 sessions (one per week for 4 weeks). Stimulation of active and latent myofascial trigger release points according to the Hong method.  Myofascial release therapy group - 4 sessions (one per week for 4 weeks). Manual therapy performed on the connective tissues according to a standardised protocol | Passive therapy |
| Ceballos-Laita | 2021 | Both groups received the same therapeutic exercise (TE) intervention, carried out by a physiotherapist. The TE program consisted of 30 sessions (3 sessions of 60 min per week for 10 weeks. Each session included an active warm-up based on low-intensity movements and dynamic stretching, a central part with aerobic training and strengthening exercises of the major muscles, and a cooldown part including static stretching and respiratory exercises.  The Pain Neurophysiology Education + TE group also received a face-to-face educational intervention This intervention consisted of 8 sessions. The PNE sessions included explanations about acute and chronic pain and the potential sustaining factors of central sensitization such as emotions, stress, illness perceptions, pain cognitions, and pain behaviour. Patients were encouraged to apply the new knowledge to their daily life. | Educational; Physical activity |
| Cedola | 2019 | 2 week anti inflammatory (ITIS) diet | Nutritional |
| Celletti | 2021 | “Felt sense" rehabilitation. With various hands on placement. Patient engages with the sense and develops cognitive adaptations to pressure and positioning. Sponges were also used to change sense of touch | Psychological |
| Cetin | 2022 | 1 hour Tai Chi 2/7 for 10wk vs. HEP of 1 hour strength, stretch and balance exercise 2/7 for 10wk | Physical activity |
| Cetin | 2020 | Tai chi group - taught each form for 1 week and combined with the previous form each week. All forms were completed at 10 weeks. Each session took 1 h (15 min for warming up exercises, 30 min of Tai Chi  forms, and 15 min for cooling down exercises).  Home exercise group - 1h home program for 2 days a week. The first and last 15 min of the exercise program consisted of  warm-up and cooling exercises. After warm-up exercises, stretching for  shoulder, hamstring and erector spinae muscles, strengthening exercises  for abdominal and back muscles were performed 10 times each for 30  min. Each of the exercise was explained and demonstrated in detail by  the experienced physiotherapist | Physical activity |
| Christie | 2015 | Participants were offered two sepa- rate periods (each lasting 12 weeks) of aquatic exercises and two 5 week periods without intervention  The aquatic exercises were performed in an indoor pool with a water temperature of at least 34°C. Each session lasted 60 minutes, including ten minutes of warm-up exercises, 15-20 minutes of resistance exercises, 20-25 minutes of aerobic exercises and ten minutes of cool-down exercises. | Physical activity |
| Cooney | 2019 | 60 minute exercise sessions, 3 x 10 min aerobic exercise at 55-85% max heart rate, plus resistance training exercises. Three sessions per week for eight weeks | Physical activity |
| Correia Moretti | 2016 | Aerobic exercise (AE) was conducted on exercise bicycle), at 50-60% of maximum heart rate monitored with a frequency meter.  Intervention group - Global, lymphatic, upper trapezius, torso, lumbar, and quadriceps pompage were applied (5 repetitions maintaining tension for 15 seconds (s) and 10 second intervals). Passive stretching of the hamstring and quadriceps femoris muscles, and active stretching of the anterior and posterior trunk muscles and the triceps muscles (two repetitions during 30 seconds and a 10 second interval).  Control group performed aerobic exercise only | Passive therapy; Physical activity |
| Coste | 2021 | Osteopathic manipulation- six sessions of 15-20 minutes, once per week over 6 consecutive weeks. Progressive mobilisation of spine, hips, shoulders, neck  Control - Sham manipulation- six sessions of 15-20 minutes, once per week over 6 consecutive weeks. Same order as treatment group but all manoeuvres stopped halfway so no significant joint manipulation and no thrust manipulation. | Passive therapy |
| Cramp | 2020 | Four group sessions and a one to one session over 12 week period, including education, support for behaviour change and a supervised exercise component. | Physical activity; Psychological |
| Cuesta-Vargas | 2011 | An individualised multimodal physiotherapy program combined with education based on cognitive behavioural principles was prescribed, delivery in groups of max 12. 60 minutes 3 times per week for 8 weeks. 15 minutes of mobility and flexibility exercises, 15 minutes of resistance and muscle strengthening exercises and 20 minutes deep water running. Individual workloads determined by pre intervention heart rate and lactic acid monitoring | Educational; Physical activity |
| Cunningham | 2019 | TEACH - 6 weekly in person sessions lasting approx 1 hour, aiming to reduce fatigue, psychological distress and pain levels. | Psychological |
| Cunningham | 2022 | Six weekly, one hour sessions of TEACH (treatment and education approach for childhood-onset lupus) cognitive behavioural therapy program and TAU | Psychological |
| Cuperus | 2015 | Face to face self management treatment program = 6 group sessions for 2-4Hr. for 6 weeks. 1x Given education about pain management and medication. 1 x exercise session, 1 x goal setting, education on graded activity for 4-6. | Educational; Physical activity |
| Da Silva | 2018 | Phototherapy group - 10 weeks, 2 sessions per week. Light sources applied on 10 tender points for 300s  Exercise training group - (phototherapy placebo) - 10 weeks, 2 sessions per week. Stretching and aerobic exercises  Phototherapy and exercise training group - 10 weeks, 2 sessions per week. Phototherapy applied 30 minutes prior to exercise session | Passive therapy; Physical activity |
| Dardin | 2022 | Two weeks of familiarisation sessions followed by 16 week resistance exercise program. Sessions consisted of three sets (10 repetitions each) at 60 to 80% of 1 repetition maximum (1RM) of the bench-supported single arm row, dumbbell side lateral raise, horizontal dumbbell chest press, unilateral forehead dumbbell triceps extension, dumbbell biceps curl, knee extension, knee flexion, hip  abduction, hip adduction, ankle plantar flexion in the leg press machine, and squats with dumbbells. | Physical activity |
| Darmant | 2021 | 2 sessions Argentinean tango per week (48 sessions) from Month 0 to month 6 | Physical activity |
| Davies | 2012 | Low GI diet group- 6 weeks. Carbohydrate intake limited to 45g/day  Calorie restricted diet group- 6 weeks. 2000kcal/day (approx 50% carbohydrates, 15% protein and 30% fat) | Nutritional |
| Demirbag | 2012 | Group 1 - Touch music aromatherapy 2/7 for 6 weeks 30-35 minutes of massage, followed by education  Group 2 - Sleep music aroma therapy 2/7 for 6 weeks 30-35 minutes sleep, followed by education | Educational; Passive therapy |
| Di Carlo | 2021 | 8 x 1/7 electroacupuncture for 16 points of the body | Passive therapy |
| Diaz-Toral | 2017 | electroacupuncture/dry needling, 2/7 for 10 weeks | Passive therapy |
| Dilekci | 2020 | All patients were given an exercise program including 15 min of standardized exercises for flexibility and stretching exercises (hamstring, pelvic and abdominal muscles).  Physical therapy (PT)group - 5/7 for 3 weeks. Applied 45C hot pack to the low back region for 20 min and 80 Hz frequency TENS for 20 min, 6 min of therapeutic continuous ultrasound. Slow circular movements were applied to the transducer head over the paravertebral low back region.  PT + Balneotherapy( BT) group - 20 min at 38-40C in thermal pools. After resting for 30 min, they received in the PT program. | Passive therapy |
| Dilekci | 2019 | Group 1 - All patients had 20 min of 45°C local heat pack and 30 min of 80 Hz frequency TENS and 6 min of therapeutic continuous ultrasound applied during each session  Group 2 - 20 min of 38-40°C mineral water bath in the curative pool in addition to the PT protocol | Passive therapy; Physical activity |
| Dobson | 2022 | Fatigue Management programme for 7 weeks in person vs online | Psychological |
| Dover | 2021 | Creatinine supplementation vs. placebo | Nutritional |
| Ducamp | 2022 | Standardised spa therapy (SST) – 72 treatments over 18 consecutive days including; individual baths in mineral water with underwater spray (36C, 10â€“20 min), mineral mud applications (40-42C, up to 15 min), massages by a registered physiotherapist (36C, up to 10 min), and collective exercises in a mineral water pool supervised by a registered physiotherapist(32-34C, 20 min).  SST + Therapeutic patient education (TPE) group attended 1.5-2 h collective educational workshops inclduding indoor sessions of stretching and relaxation exercises adapted to patients with fibromyalgia, and three collective outdoor sessions of adapted aerobic activity | Educational; Passive therapy; Physical activity |
| Durcan | 2014 | A 12-week home exercise program was prescribed for the intervention group. Participants were assessed by a doctor (LD) and senior physiotherapist (FW) at baseline and then every 3 weeks for the duration of the program.  a walking program was devised according to ACSM guidelines on physical activity. Participants were given a step count target and advised on level of exertion to aim for. Resistance exercise to be done 3x weekly and range of motion exercises to be done daily | Physical activity |
| Dures | 2022 | 1:1 face to face session x 4 | Psychological |
| Durmus | 2009 | 20 exercise home exercise program (HEP) 7/7 for 12-weeks. Exercises were taught first by physio. they were called weekly by the researcher and checked whether they were performing the program or not | Physical activity |
| Elefante | 2022 | Physical exercise sessions were carried out using a digital platform. Patients were asked to participate in at least 70% of the lessons. The physical exercise program included moderate intensity aerobic exercises (muscle strengthening, joint mobility, breathing, static and dynamic stretching, balance and neuro-dynamics); workouts were performed 3 times a week, consisting of 60 minutes each. The program lasted for 12 weeks. | Physical activity |
| Elnaggar | 2022 | Standard exercise program - 40 minutes, three times per week for 3 months. 5 minute warm, 30 minutes exercise loading (flexibility, strengthening and weigh-bearing), 5 min cool down  Standard exercise program plus laser light therapy (LLT). LLT delivered using handheld device directly to the skin at 8 points for 30 seconds | Passive therapy |
| Elnagger | 2021 | Aqua-plyo, 3/7 for 4 weeks. 45 minutes of warm-up, exercises and cool down | Physical activity |
| Ericsson | 2016 | Person-centred progressive resistance training, group sessions. The resistance exercise was initiated at 40 % of 1 repetition maximum (RM) and progressed up to 80 % of 1 RM during the 15 weeks. | Physical activity |
| Etnier | 2009 | 60 minute sessions 3 times weekly for 18 weeks. During the exercise sessions, participants walked, performed light resistance exercises, and performed static-bridging and stretching exercises. | Physical activity |
| Evans | 2013 | 6 weeks of twice weekly classes, 1.5 hours in duration. A working list of yoga poses was developed by a senior yoga teacher, using props and addressing individual limitations as required. | Physical activity |
| Feldthusen | 2016 | 12-week person-centred physical therapy. A self-care plan was jointly developed and focused on tailoring health-enhancing physical activity and balancing life activities. | Physical activity |
| Ferwerda | 2017 | internet-based CBT tailored to individual Ieeds. Completed at least 1/4 modules Each treatment module consisted of several assignments and psychoeducational texts. with regard to pain, fatigue, negative mood, and social functioning. Lasting between 9 and 65 weeks | Psychological |
| Fjerstad | 2017 | 6-9 sessions of cognitive therapy sessions, in addition to treatment | Psychological |
| Fontaine | 2010 | Group 1 - lifestyle physical activity TAU plus 6 x 60minute group sessions over 12 weeks. based on “active living every day” - a cognitive-behavioural physical activfty programme.  Group 2 - Fibromyalgia education - 3 x monthly meetings, minimal education 45-60 minutes. Q&A session (20 minutes), social support 20 minutes. | Educational; Physical activity; Psychological |
| Frade | 2023 | 2/7 x 8weeks.  45 minutes supervised telehealth programme online. Moderate intensity, 30 minute strength circuit with progressions over the duration | Physical activity |
| Fritsch | 2022 | 3 weeks Balneotherapy. 15 sessions 30 minutes each. | Passive therapy |
| Fuchs | 2013 | Self investigation using SCM then 6 weekly 1 hour sessions of psychological intervention - Adolescents assess daily situations and how they cope with them. Guided by the counsellor, the adolescents are encouraged to gradually move from assessment to change. The adolescents are stimulated to create and engage in novel situations, to develop and reorganize their personal narratives. | Psychological |
| Gavilan-Carrera | 2023 | LBE group - 24 weeks land-based exercise protocol 3/7 45-60 mins warm-up (8-10 min), conditioning [muscle-strengthening (15-17 min) and aerobic (15-25 min)], and cool-down (10 min).  WBE group - 24 weeks Water-based exercise protocol 3/7 45-60 mins warm-up (8-10 min), conditioning [muscle-strengthening (15-17 min) and aerobic (15-25 min)], and cool-down (10 min). | Physical activity |
| Gavilan-Carrera | 2022 | Group session 2/7, 75minutes of moderate to vigorous continuous aerobic treadmill routine. for weeks 1-4 then added interval exercises weeks 6-8 and then did interval only for weeks 9-12 | Physical activity |
| Giannotti | 2014 | Rehabilitation protocol - educational-behavioural program and physical exercise. 2 days per week for 20 sessions, approx 1 hour long. Conducted as a supervised group | Educational; Physical activity; Psychological |
| Gizinska | 2015 | Whole body cryotherapy group - They passed through pre chambers (-10C and -60C) into the therapy-chamber (-110C), where they stayed for 3 min, walking in a circles and performing energetic movements with the upper limbs. Daily treatments for 2 weeks  Traditional rehabilitation group - included magnetotherapy, electrotherapy, ultrasound therapy, and laser therapy. The type of treatment and the dose were individually tailored for each patient. Daily treatments for 2 weeks | Passive therapy; Physical activity |
| Gok Metin | 2016 | Aromatherapy massage 15 minutes on each knee, 3 times weekly for 6 weeks vs. Reflexology applied for 20 minutes on each foot, once weekly for 6 weeks | Passive therapy |
| Gomez-Centano | 2020 | Nutritional supplement containing coenzyme Q10, magnesium and tryptophan for 3 months | Nutritional |
| Greco | 2008 | Group 1 - Acupuncture (AC) - TAU + 10 sessions over 5 weeks. Needles inserted to a depth sufficient to produce a needling sensation, stimulating specific points  Group 2 - Minimal needling (MN) - TAU + 10 sessions over 5 weeks. Minimal needling involves shallow insertion of needles on body areas that are not known to correspond to AC points | Passive therapy |
| Groessl | 2020 | Usual care + yoga - 2 60 min sessions per week for 12 weeks. Hatha yoga of mild-moderate intensity, designed to be safe for cLBP and accommodate adaptations | Physical activity |
| Gulsen | 2022 | Exercise group - monitored sessions attended twice a week for 8 weeks. Combined exercise training with 30 mins aerobic training and 30 mins pilates, ten minute rest in between.  Exercise + immersive virtual reality (IVR) group - monitored sessions attended twice a week for 8 weeks. Combined exercise training with 30 mins aerobic training, 30 mins pilates and 20 mins IVR treatment. Ten minute rest in between. | Physical activity |
| Gunendi | 2008 | Electrotherapy and thermotherapy vs. Electrotherapy and thermotherapy plus 30 minute treadmill 5/7 for 4 weeks | Physical activity |
| Gyulai | 2015 | Group 1 - Standard complex physical therapy + Bio-Electro-Magnetic-Energy-Regulation (BEMER); physiotherapy plus 20 min active BEMER session  Group 2 - Standard complex physical therapy + placebo BEMER; physiotherapy including individual and group exercises (30 minutes), underwater whirlpool massage (10 minutes), TENS therapy on the low back or knee (15 minutes every day), and aquagym (30 minutes every other day). 20 minute placebo BEMER session | Passive therapy |
| Hackett | 2016 | Fatigue CRESTA clinic - assessment by physician and occupational therapist then offered individualised therapy which may include OT, PT, CBT or a combination of therapies. | Physical activity; Psychological |
| Hadianfard | 2012 | Acupuncture - three sessions per week for 6 weeks vs standard care - fluoexetine 20mg daily for 8 weeks | Passive therapy |
| Hakimi | 2020 | Rehabilitation programme over 9 weeks). 3 days per week. Activity time divided across physical activities and education and wellbeing. Activities included balneotherapy, ergometer exercises, occupational therapy, physical activity, physiotherapy, walking, proprioception exercises, sophrology, yoga exercises and patient education workshops. | Educational; Physical activity; Psychological |
| Hakimi | 2023 | 9 week rehabilitation program (RP) 2 days per week for 4 weeks, 1 week rest, 2 days per week for 4 weeks. Daily activities consisted of 4 one-hour workshops including occupational therapy, physiotherapy, sophrology, various physical activities focused on muscular endurance, coordination, balance, and proprioception, and therapeutic patient education workshops | Educational; Physical activity; Psychological |
| Hammond | 2008 | Group 1- Modular cognitive behavioural approach program - two modules each with four 2.5hr meetings and a 2h review meeting over a 3-9 month period. Each meeting included self-monitoring, skills training with individualized feedback and advice, goal-setting and action planning to follow individually determined home activity and exercise programmes working towards recommended frequency targets.  Group2- information focused (standard) program - 5 2 hr group meetings | Educational; Passive therapy; Physical activity; Psychological |
| Harper | 2021 | Participants provided with a PA and behavioural change support programme and a wrist-worn activity tracker in addition to standard care. Week 1-8 was weekly face to face PA sessions in groups and phone calls to support behaviour change and motivation. Weeks 9-12 were weekly phone calls only. | Physical activity; Psychological |
| Hasni | 2022 | aerobic sessions 3/7 for 12 weeks, goal 30 minutes vigorous exercise | Educational; Physical activity |
| Haugmark | 2021 | Vitality Training Program (VTP) - 10 weekly 4 hour sessions and a booster session after 6 months. Participants explored a range of topics using creative methods, attended mindfulness meditation and yoga. | Educational; Physical activity; Psychological |
| Hays | 2018 | 3 months of chiropractic treatment | Passive therapy |
| Hennard | 2011 | yoga and meditation | Physical activity |
| Hergenroeder | 2022 | Group 1 - 6months. behavioural counselling, sit-stand desk, initial consult followed by 5 monthly phone calls  Group 2 - completed online survey at T0 and T6 months and offered a 60-minute review at the end of the study | Educational; Psychological |
| Hewlett | 2011 | CBT intervention, 2 hours per week covering key topics such as fatigue validation, energy management, sleep, goal setting, stress and relaxation, assertiveness, self-help toolkit, dealing with setbacks. Use of reflective questioning, goal setting, problem solving and self monitoring.  usual care group - 1 hour information session delivered by rheumatology specialist nurse | Psychological |
| Hewlett | 2019 | 7 CBT sessions 2hrs/wk. for wk 1-6 and then a consolidation session in week 14. Tutors use exploratory questioning, goal-setting and peer-support to enhance self-efficacy | Psychological |
| Higgins | 2019 | Access to an online PAIN EASE programme - self-guided, CBT programme, pedometer. weekly phone calls | Psychological |
| Holla | 2009 | Participation within the centres activity options. These are capability specific exercise offered to members upon joining. | Physical activity |
| Hoon | 2017 | Take Charge of Pain programme, TCOP. 2 x 2.5hr sessions over 2 weeks. explore the benefits and barriers to healthy eating and physical activity, managing medication, developing meaningful networks, positive thinking and challenging unhelpful thoughts, and dealing with flare-ups. | Educational |
| Houghton | 2018 | Home based exercise intervention - home visit to individually tailor program to participant abilities, provided with pictures of exercise, schedule for progression, therabands, hand gripper and modified exercises in case of joint pain. 26 weeks, six 4 week blocks of jumping exercises (3sessions/ week) followed by 2 weeks of resistance training with therabands and handgrips. Additionally one group session/month | Physical activity |
| Hsieh | 2014 | Intervention - Active light therapy 3 x 40-minute treatments weekly for 2 weeks  Placebo - all participants received three 40-min hot-pack treatments weekly for 2 weeks. The light device was used for all the participants, but electrical power was supplied to the unit for the treatment group only | Passive therapy |
| Huang | 2019 | Conventional therapy plus a brief Guandong Online Hospital (GOH) intervention including education, medical suggestions and psychotherapy | Educational; Psychological |
| Husakova | 2019 | 6-month long intervention consisting of twice weekly group supervised physiotherapy. The exercise unit consisted of three sections: warm-up with cardiorespiratory fitness, the main session focused on the practice of correct activation of deep core stabilisation and involvement muscles in motion, spinal traction and balance training, and cool-down with relaxation. | Physical activity |
| Ide | 2008 | Group 1 - 1hr supervised activities such as card games and music etc. no PA., 1/7 for 4 weeks plus:  aquatic respiratory exercise-based program, in a 32C heated pool, 1.05 m deep, for 1 h in the evening, four times a week, for 4 weeks.  Group 2 - TAU and 1hr supervised activities such as card games and music etc. no PA., 1/7 for 4 weeks | Physical activity |
| Inderjeeth | 2021 | weekly 2.5 hr SM education session for 6 weeks, including stretches and supervised exercise classes | Educational; Physical activity |
| Jastrzabek | 2013 | Group 1 - 10 days twice a day received 3-min liquid nitrogen flow treatment, with the gas temperature at the nozzle approximately -160 C. Each patient performed physical exercises according to their condition under the supervision of physiotherapists and continued conventional medical treatment for RA.  Group 2 –10 days twice a day received 3-min cool air blowing treatment. The air temperature at the nozzle was approximately -30 C. Each patient performed physical exercises according to their condition under the supervision of physiotherapists and continued conventional medical treatment for RA. | Passive therapy; Physical activity |
| Jiao | 2019 | Guided by a physician, the participants  met for 1 hour twice weekly, for 12 weeks. Each participant  was given an instructional handout and video showing an instructor performing Ba-Duan-Jin. All subsequent sessions, each led by a physician, began  with a brief review of the Ba-Duan-Jin movements, breathing and relaxation  techniques, followed by a warm-up session leading into repetition of the eight  forms of Ba-Duan-Jin. Encouraged to do daily practice of 16 mins at home | Physical activity |
| Jones | 2023 | EnhanceFitness (EF) programme 3/7 for 12 weeks, strength, stretch and balance. | Physical activity |
| Kabiri | 2018 | All 3 groups underwent routine physiotherapy using TENS and FARADIC (Iran) at 50 Hz current.  Group 1 - Massage – after physiotherapy, classical massage was done on the knee. Sweet almond oil to reduce friction of the area. The amount of oil used in each session  was 3 mL and massage therapy was conducted within 10 sessions, each session lasting 20 minutes, and three sessions per week  Group 2 - Aromatherapy - two drops of lavender essential oil was embedded in a cotton ball and then the cotton ball was attached to the collar from 10:00 pm to 7:00 am at home every other day for one month  Group 3 - TAU | Passive therapy |
| Kankaya | 2020 | Online education website - resources for each new topic uploaded weekly and questions were answred through the website. After 3 months participants were informed by telephone that the education was over but individual counselling and information updates could continue as required. | Educational; Psychological |
| KARACA | 2022 | Initiation of anti-TNF therapy plus exercise therapy to include the BETY (Cognitive Exercise Therapy Approach). Patients participated in group exercise session for 12 weeks, 3 times per week in supervised sessions lasting approximately 1 hr | Physical activity; Psychological |
| Karkon | 2023 | Group 1 - received a Fatigue and Activity Management Education for Work (FAME-W) handbook  Group 2 - four week online FAME-W training in addition to handbook - group based self management, 1.5h per session | Educational |
| Karper | 2016 | Ongoing exercise programme, three times per week for 11-12 months per year. Centred on activities that progressively develop walking endurance, muscle strength, and endurance in major muscle groups and range-of-motion in joints and muscles using mild- to moderate-intensity activity | Educational; Physical activity |
| Katz | 2018 | Education only (EDUC). The EDUC group received an educational brochure (Be Active Your Way: A Guide for Adults [http://health.gov/paguidelines/pdf/ paguide.pdf]) and a guided discussion of simple ways to increase physical activity in daily life based on the booklet.  Pedometer only (PED). The PED group received the educational booklet and discussion, plus a pedometer and a diary to record daily step counts from the pedometer. Participants were shown how to wear the pedometer, and were instructed to wear it from the time they got out of bed in the morning until they went to bed at night,  Pedometer + step targets (PED+). The PED+ group received the educational booklet and discussion, the pedometer and step diary, and individualized daily step targets. Step targets were based on the week of activity monitoring between the baseline and randomization visits, and were calculated to increase participants average daily step counts by 10% for every 2 weeks of the intervention period | Educational; Physical activity |
| Keramiotou | 2020 | Control group had four sessions of training in alternative methods of performing daily activities, use of aids, joint protection and energy  conservation,  Exercise group received by the hand therapist a 30-min daily programme at home of strengthening and stretching upper limb exercises for  12 weeks, in addition to routine care. The programme included 9 strengthening and stretching exercises for the upper extremities with a stick, 10 strengthening and stretching exercises for the fingers and 11 strengthening exercises against resistance with therapeutic putty | Physical activity |
| Kilic | 2020 | Progressive Muscle Relaxation technique, The PMR and training were carried out in a private room with sufficient light and ambient temperature to make the patients feel comfortable. A CD of PMR was given to the patients that explained how certain muscles in the body should be contracted and relaxed. The muscle groups used for the PMR were those of the toes, feet, legs, calves, buttocks, thighs, abdominal muscles, back muscles, chest, hands, biceps, triceps, shoulders, neck, face and tongue | Physical activity |
| Kim | 2023 | Heat massage using a mechanical device. Rollers applied heat and massage while automatically adjusting to the curvature of the length of the spine. All participants used the device for 40min, once daily, 5 days per week for 4 weeks.  Standard care; physical therapy group - 5 min therapeutic ultrasound, 10 min TENS, 15 min hot pack application. | Passive therapy |
| Klingberg | 2019 | Very Low Energy Diet (VLED) 640kcal/day consisting of four daily portions of powder consumed as shakes or soups. Followed for 12 or 16 weeks depending on initial BMI. Food was then reintroduced over 12 weeks. Participants were given a personalised energy target based on individual energy requirments, reduced by 30% to achieve further weight loss. Treatment given within 12 months and supported by medical follow up. | Nutritional |
| Knittle | 2015 | Intervention; week 1 - small group educational session and signposting to information resources and local classes. Week 2 - 1:1 motivational interview including goal setting and introduction of exercise diaries Weeks 4&5 - two 1:1 self regulation coaching sessions Weeks 6,12&18 - follow up phone call | Educational; Physical activity; Psychological |
| Kocyigit | 2016 | Group 1 - FMS education given at beginning of treatment, day 15, then after 1 month, 3 months and 6 months  Group 2 - FMS education as above and then SPA treatment for 20 minutes 5/7. Organised between 9-11am, in mineral water at 34.8C. 21 sessions in total, supervised | Educational; Passive therapy |
| Kong | 2021 | 8 x 90minutes CBT-based therapy over 8 weeks | Psychological |
| Kucharski | 2019 | A gym-based exercise programme with physio 3/7 for 20 weeks. exercise consisted of moderate-to- high intensity aerobic and resistance exercise with a total of 27 min of aerobic exercise and 5 resistance exercises for large muscle groups  Active control - light home-based exercise for mobility, lower body strength and balance | Physical activity |
| Lami | 2016 | Males received CBT-I from male psychologists, group sessions 1/7 for 9 weeks. Each 90 minutes. Given a manual for home use.  Females got CBT-I from female psychologists group sessions 1/7 for 9 weeks. Each 90 minutes. Given a manual for home use. | Psychological |
| Lami | 2018 | CBT- insomnia & Pain - Focussed on sleep hygiene rules, relaxation , planning for rest  CBT -Pain - focussed o nemotional responses to stress and other states | Psychological |
| Latocha | 2022 | CBT-Insomnia - 6 weeks group sessions face to face | Psychological |
| Lazaridou | 2019 | 6-week yoga programme, Satyananda Yoga is a traditional form of yoga which includes asanas (physical exercises), pranayama (breathing exercises), pratyahara (mind focusing practices), and meditation (breathing awareness, awareness of senses, awareness of thoughts, and yoga nidra). 1.5hrs / week, 30-min daily Video recording given to allow participants to practice at home | Physical activity |
| Li | 2020 | 8-week intervention included 3 components: 1) an in-person session with 20 minutes of group education and 30 minutes of individual counselling with a physical therapist, 2) use of a Fitbit Flex 2 with account access in the company web-site, and 3) 4 biweekly phone calls (20-30 minutes) from a physical therapist. | Educational; Physical activity; Psychological |
| Lin | 2020 | Participants in both groups had hot packs applied to both knees for 20 mins followed by TENS for 20 mins.  Video games treatment; Followed by 20 minutes video games with two sessions that required participants to move their trunk and lower limbs as quickly as possible on a step-sensing pad.  Therapeutic exercise group; Stretching warm up, cycling for 10 mins at approx 40-60% max HR and a stretching cool down | Physical activity |
| Lindqvist | 2018 | Intervention - 20 nutritionally balanced vegetarian dishes plus 20 portions of blue mussel (to be combined by participant) delivered every 4 weeks. Consumed one meal daily, 5 days per week for 11 weeks total. | Nutritional |
| Liu | 2012 | Qigong, 2 training sessions, then 2 x per day at home, 1/7 in group class. for 6 weeks. vs. sham Qigong exercise was designed to match the body movements of the Qigong exercise in frequency and intensity. However, no meditation or healing sounds were involved in the sham Qigong exercise | Physical activity |
| Ljotsson | 2014 | Treatment was delivered over 10 weeks. Tasks and information were presented and participants were encouraged to contact therapists at least once a week to discuss their work. Treatment steps were sequential and participants were encourage to complete the first 4 (of 5) in the first half of the treatment period. | Psychological |
| Loeppenthin | 2022 | The intervention consisted of 18 intermittent aerobic exercise training sessions: 3 sessions of 20 to 30 minutes per week for 6 weeks. The sessions were performed on bicycle ergometers (Kettler) | Physical activity |
| Lopes-Souza | 2021 | The WBVE group performed training program twice a week with at least one day between each session for 12 weeks. In the first 4 weeks, WBVE session consisted of 10 bouts with 30 s performed within a frequency of 30Hz, D 1.23 mm and a peak of 2.22g. From 5 to 8 week, WBVE session consisted of 10 bouts with 60 s performed within a frequency of 40Hz,D0.95mm and a peak of 3.06g. From 9 to 12 week a WBVE session consisted of 10 bouts with 60 s performed within a frequency of 50Hz, D 0.88 mm and a peak of 4.40g.  Isometry group - the same warm up was performed by both groups and they adopted the same stance on the vibration plate, but it was not switched on for the isometry group. | Passive therapy |
| Lorig | 2008 | Web based instruction (The Learning Centre) and bulletin board discussion (The Discussion Centre), a personal toolkit including exercise and medication diaries and tailored exercise programs (My Tools) and the Arthritis Helpbook.  Delivered over 6 weeks asked to log in 3+ times per week to read the weekly content, post an action plan and participate in self tests. Can also respond to questions/problems raised by other participants | Educational; Physical activity; Psychological |
| Luciano | 2011 | Usual care from GP and a psychoeducative program. Nine 2hr sessions over 2 month. 5 education sessions and 4 autogenic training sessions | Educational |
| Lumley | 2016 | Emotional Awareness and Expression Therapy (EAET) - all groups had a 90 minute session once a week for 8 weeks  CBT - all groups had a 90 minute session once a week for 8 weeks  Active education control condition - all groups had a 90 minute session once a week for 8 weeks | Psychological |
| Maarj | 2023 | Following a 3D scan of the feet, participants were fitted with custom-made orthotics and given verbal/written instructions on how to gradually increase wear time. | Passive therapy |
| Maddali Bongi | 2016 | Tai Ji Quan (TJQ) practice for 60 min sessions, twice per week for 16 weeks. Exercises tailored to clinical situagtion of each participant. Also daily home exercise for 15 min twice per day using a DVD | Physical activity |
| Maddali-Bongi | 2011 | Body Movement and Perception (BMP) method - group gymnastics based on awareness of body perception, low impact pjhysical exercise and relaxation. 2x 50min session per week | Physical activity; Psychological |
| Maddali-Bongi | 2019 | Mind body therapy (MBSR) - weekly sessions lasting 2.5-3h for 8 weeks and one all day intensive class. Mindfulness activities are practiced in class and as homework | Psychological |
| Maindet | 2021 | 3wk spa therapy, 6/7 (18 days of Treatment). 1. Hydromassage baths 2. Hydro-mineral mud applications 3. Body jet showers with adjustment of the intensity of jets 4. Water affusion massages 5. Collective exercise in a mineral water pool under supervision of a state-registered physiotherapist They also had conferences and walking therapies. | Educational; Passive therapy |
| Martin | 2014 | The same treatment team managed all of the groups. Each patient attended twice-weekly group sessions of 105 minutes, for 6 weeks (a total of 12 sessions). each week, one of the sessions consisted of 1 hour with the psychologist plus 45 minutes of educational activities with a physician and a psychologist another session included 1 hour with a psychologist plus 45 minutes with a physiotherapist. | Educational; Physical activity; Psychological |
| Martinez | 2014 | Group 1 CBT-I: Session 1 was focused on providing information about the relationship between sleep and FM, In Session 2, participants were given instructions for applying sleep restriction and stimulus control. Session 3 was aimed at training physiological deactivation procedures (slow breathing, passive relaxation and imagery training). Sessions 4 and 5 were focused on cognitive therapy to change negative thoughts about insomnia through verbal discussion and behavioural experiments. Session 6 was devoted to maintaining achievements and preventing relapses  Group 2 - Sleep Hygiene: In Session 1, participants were given the same information about sleep as those in the CBT-I program. Session 2 was devoted to sleep hygiene rules related to environmental factors (e.g., noise, temperature, light). Session 3 was focused on learning about lifestyle factors that influence sleep (use of stimulants and other substances). Sessions 4 and 5 were aimed at providing information about diet and physical exercise, respectively. Session 6 was devoted to maintaining achievements and preventing relapses, as in the CBT-I program. | Educational; Psychological |
| McCormack | 2018 | FAME-W is an occupational therapy-led, 4 week, group-based self- management programme focused on managing fatigue in the workplace, with participants attending once per week for 2 h. Each sessions includes education and individual goal setting. Workplace visits were offered but no participants accepted this. | Educational |
| McCormick | 2022 | Web based acceptance and commitment therapy (ACT) program. 2 x 1hr webinars, 1 per week for 2 weeks. Each session included 40 mins ACT based education followed by 20min Q&A session | Psychological |
| McCurry | 2021 | CBT for insomnia - sleep hygiene education, cognitive strategies and in-bed restriction plan  Education only control - designed to control for non-specific treatment effects (social support, education) | Educational; Psychological |
| Meenakshi Sundaram | 2015 | Steam therapy and functional rehabilitation vs Whole body cryotherapy and functional rehabilitation | Passive therapy |
| Mete | 2019 | Core stabilisation exercise - 2 days per week for 6 weeks vs Combined exercise - 2 days per week for 6 weeks | Physical activity |
| Metri | 2023 | 60 min yoga session, four times per week for 6 week, held at workplace (school) after work hours | Physical activity |
| Mhalla | 2011 | Transcranial Magnetic Simulation (TMS) 1wk baseline obs. The treatment protocol consisted of a total of 14 stimulation sessions over 21 weeks and one follow-up visit at week 25. Patients were seated in a comfortable reclining chair and asked to keep their hands as relaxed as possible. Magnetic stimulation was applied with a MagPROX100 machine, using a figure-8-shaped coil oriented at a tangent to the scalp, with the main phase of the induced current in the anterior-posterior direction. The patients were fitted with ear plugs during TMS. | Passive therapy |
| Miro | 2022 | The app includes 9 modules that provide information about FMS and its treatment (i.e., General introduction and treatment objectives; Pain; Coping skills; Stress and relaxation; Medication use; Sleep; Physical activity; Thought management; and Relapse prevention. Each module has different resources which are accessible after they are unlocked. The treatment unfolds over the course of 47 treatment days. | Psychological |
| Mishra | 2014 | Acupuncture vs. simulated acupuncture | Passive therapy |
| Mist | 2018 | Group 1 - Twenty treatments of acupuncture, 2/7, using traditional chinese medicine (TCM) diagnosis based point selection  Group 2 - group education, facilitated discussions about diagnosis and treatment | Educational; Passive therapy |
| Miyamoto | 2019 | Walking exercise , supervised, outdoor track 3/7 for 16 weeks. | Physical activity |
| Moe | 2016 | new MDT approach, received 3.5hr group educational programme followed by individual consult with rheumatologist and MDT personnel as required (orthopaedic surgeon, physical therapist, occupational therapist, pharmacist, or dietitian) | Educational |
| Moreira | 2023 | 2/7 for 6 weeks 20 minutes each session. Traditional Chinese medicine acupuncture and auricular (French) acupuncture | Passive therapy |
| Morsley | 2017 | 6 week Progressive Resistance Training (PRT), groups of up to 10 - Tailored to individual where needed, resistance and reps increased over the 6 weeks. 1/7 for 30-60 minutes. Encouraged to exercise 2/7 at home with an illustrated sheet for support. | Physical activity |
| Murphy | 2010 | Group 1 - tailored activity pacing. received a study-specific education module on activity pacing. he OT tailored recommendations based a personalized report that detailed the relationship between activity and symptoms using graphs and bulleted points.  Group 2 - general activity pacing. received a study-specific education module on activity pacing. In the general instruction group, the OT discussed the general principles of activity pacing (i.e., pre-planning activities, alternating activity with rest before a symptom exacerbation) with a recommendation to implement the strategies over the subsequent few days. | Educational |
| Murphy | 2016 | General activity pacing intervention - care 3x 1:1 sessions with OT. between 30-60 minutes each time. about 7-10 apart, wore an actiwatch  Tailored activity pacing intervention - care 3x 1:1 sessions with OT. between 30-60 minutes each time. about 7-10 apart. Then received an activity report based on data collected at home from the actiwatch. This was then discussed at each session with the aim of improving responses. | Physical activity |
| Murphy | 2023 | Resilience building energy management to enhance wellbeing (RENEW) - 12 week online peer-supported self management program.. Nine health coaching sessions online, weekly for first 6 weeks, then biweekly for six weeks. 15-30 mins | Educational; Psychological |
| Nadal-Nicolas | 2020 | Two sessions per week for four weeks. manual therapy: massage to posterior cervical muscles. | Passive therapy |
| Ndosi | 2013 | Rheumatologist led care (RLC) - TAU. Approx 15 minute care slot, takes history, performs physical examination, pain control, prescribing or recommending medication and dosage changes, intra-articular or intramuscular steroid injections, provision of patient education, psychosocial support and ordering blood tests or X-rays. Referrals for ward admission or other health professionals, were carried out as appropriate  Nurse led care (NLC). Approx 30 minute care slot, takes history, performs physical examination, pain control, prescribing or recommending medication and dosage changes, intra-articular or intramuscular steroid injections, provision of patient education, psychosocial support and ordering blood tests or X-rays. Referrals for ward admission, to the rheumatologist or other health professionals, were carried out as appropriate | Passive therapy |
| Neuberger | 2007 | class exercise in a fitness centre 12 weeks low impact exercises for 1hr, 3/7. Vs home exercise via a video tape. same ex rx in both groups | Physical activity |
| Neudorf | 2021 | YMCA move for health exercise programme for 12 weeks | Physical activity |
| Niedermann | 2013 | Standard flexibility exercise - all study participants received the current standard of care, a weekly 1-hour exercise group supervised by a physiotherapist, with focus on spinal flexibility.  Cardiovascular training and flexibility exercise. The training group performed a 12-week supervised nordic walking training for 30 minutes twice a week using individually monitored, moderate-intensity heart rate (HR) levels.  Control (attention control and flexibility exercises) The attention control intervention consisting of monthly 2.5-hour discussion groups on coping strategies and techniques of mindfulness-based stress reduction led by a psychologist. | Physical activity |
| O'Connor | 2013 | 3 days of hydrotherapy | Passive therapy |
| Oksuz | 2023 | Group 1 - Aerobic Training 3/7, warm up, treadmill for 20 minutes at increasing HRM from 40% to 75%. for 6 weeks, and then the last 2 weeks HRM was 60-75% and 30 minutes.  Group 2 - Aerobic Training as above plus.   plus Clinical pilates exercises. | Physical activity |
| Oosterveld | 2009 | Infrared sauna - The temperature in the cabin can be adjusted from normal ambient room temperature up to 90C. The patients were seated in the infrared cabin, which has six heating sources; three at the back, two in front besides the entrance, and one under the bench behind the lower legs of the patients. The infrared used has a long wavelength between 5,000 and 1,000,000 nm. The patients were treated for a period of 4 weeks, twice weekly, with eight IR sessions in the IR cabin (30 min at an ambient temperature of 55C). | Passive therapy |
| O'Riordan | 2017 | Fatigue and Activity Management Education (FAME) - 2.5h session once per week for 6 weeks, divided equally between group education and individual goal setting | Educational; Psychological |
| Otter | 2010 | 45 minute reflexology treatment, weekly for 6 weeks vs.45 minute foot massage, weekly for 6 weeks | Passive therapy |
| Ozcadirci | 2019 | Biopsychosocial exercise approach (BETY) - one hour, three times per week for 12 weeks. Included clinical pilates, dance therapy-authentic movement and pain management information. | Educational; Physical activity |
| Ozyasar | 2018 | Supervised aerobic exercise vs. Unsupervised aerobic exercise | Physical activity |
| Paek | 2018 | Education using RA T2T strategy including monthly telephone counselling for motivation and 4 individualised face to face education sessions at baseline, 3,6 and 9 months | Educational |
| Palekar | 2014 | Pilataes vs Yogasanas, 2 weeks | Physical activity |
| Park | 2017 | Sit 'N' Fit Chair Yoga - twice weekly 45 min session for 8 weeks. Post intervention participants were given a handbook and sked to report yoga home practice for following three months  Health Education Group (attention control group) - twice weekly 45 min education sessions for 8 weeks. | Physical activity |
| Passard | 2007 | repetitive transcranial magnetic stimulation | Passive therapy |
| Patru | 2021 | Group 1 - CBT and OT, 12 sessions 1/7 for 2 hours not described  Group 2 - Kinetic therapy: The kinetic therapy intervention included four  types of sessions: Aerobic exercise and balance, postural hygiene,  stretching and pilates. Patients attended one session per week at  the gym and were recommended to try to perform the exercises  at home two more times a week, | Physical activity; Psychological |
| Polat | 2021 | Conventional Training Group (CTG) - aerobic exercise program that comprised cycling activity for 20 minutes 3 days per week. Aerobic exercise intensity was adjusted to generate heart rates equivalent to 60%â€“70% of age-adjusted maximum heart rates.  Virtual Reality Group (VRG) - aerobic exercise program that comprised cycling activity for 20 minutes 3 days per week. Aerobic exercise intensity was adjusted to generate heart rates equivalent to 60%â€“70% of age-adjusted maximum heart rates | Physical activity |
| Poole | 2013 | Participants were sent the workbook and DVD and expected to complete the program in 3 to 4 months. Each chapter of the workbook contained learning activities, with action plans modelled after arthritis self-management programs. The DVD demonstrated face, mouth, hand, arm and leg exercises. | Educational; Physical activity |
| Poole | 2014 | Access to the interactive education format for SSC . The program had 10 modules, an exercise video, worksheets and resources. Message board for questions | Educational; Physical activity |
| Pot-Vaucel | 2016 | stage 1: private interview with a therapeutic education nurse, used to make an educational diagnosis and, with the patient, choose 3 subjects which cause problems, out of the 10 suggested; â€¢ Stage 2: allocation to an educational route leading in 6 months to communal workshops (maximum of 3) and/or private interviews for the 3 subjects chosen; â€¢ Stage 3: final interview with a therapeutic education nurse spe- cialising in training evaluation, leading to production of a report.   The patients had the choice between the possibility of individual treatment (social worker, occupational therapy, dietician, psychologist, chiropodist) and joint workshops: â€¢ understand my disease and talk about it (presented by a rheumatologist and a PTE instructor); â€¢ living with my treatment (presented by a rheumatologist and a PTE instructor); â€¢ maintain my mobility and save my joints (presented by an occupational therapist and a physiotherapist); â€¢ identify my rights (presented by a social worker and a PTE instructor); â€¢ Diet: between balance and pleasure (presented by a dietician and a PTE instructor); â€¢ self-image, emotion and morale (presented by a psychologist and a PTE instructor). | Educational |
| Primdahl | 2014 | Control - rheumatologist group. Participants in all groups completed two 3h multidisciplinary session to enhance self-efficacy and knowledge of how and when to seek help. Follow up as usual with rheumatologist every 3-12 months based on rheumatologist judgement.  Shared care group. No planned consultations apart form annual hospital reviews. Could address problems to either GP or to specialist rheumatology nurse-led helpline. GPs could contact department and use helpline for specialist advice and review  Nursing group. Thirty min consultation every 3 months with a rheumatology nurse and could address concerns to nurse helpline. Seen by rheumatologist if DAS28 exceeded 3.2. Nurse could consult rheumatologists and make referrals to GP, OT, physiotherapist or podiatrist. | Passive therapy |
| Prioreschi | 2016 | Whole body vibration therapy consisted of two 15 minute session per week (total of 24 sessions over 12 weeks) of supervised therapy, which comprised of ten repetitions standing on the vibration plate for 60 seconds, followed by a 30 second rest period | Passive therapy |
| Puksic | 2021 | The yoga program was conducted two times weekly with 90 min per  session for 12 weeks at the Departments gym. The program was performed accord ing to the Yoga in Daily Life system (Level 1 - Sarva hita asanas)  The educational control group had once weekly 60 min lecture by a  rheumatologist on arthritis-related topics (e.g. inflammatory process  and joints, RA symptoms, comorbidities, therapy and treat-to-target  approach, exercise and self-help) with participant discussion included. | Educational; Physical activity |
| Racine | 2019 | Operant learning (OL) - one 2hr session weekly for 10 weeks. The OL approach aims to reinforce a switch from pain-contingent to goal-directed behaviour using activity-rest cycling or moving at a â€œslow and steady paceâ€ to achieve daily life activities/goals.  energy conservation (EC) 10 x 1/7 for 2 hours. the EC perspective focuses on patient energy expenditure and seeks to achieve a balance between accomplishing day-to-day valued activities/goals and resting to reduce pain and fatigue. | Educational |
| Rapoliene | 2020 | Control group did not have a bath. Tap water group - 400L bath at 36C for 20 minutes, as per mineral bath groups  IG1 – 20 g/L bath was filled with 73 L of mineral water and 327 L of pure tap water. Bath for 20minutes, soft towel dry and no shower for at least 1-hour post immersion 5/7 for 2 weeks (10 baths).  IG2 – 40 g/L was filled with 145 L of mineral and 254 L of pure tap water Bath for 20minutes, soft towel dry and no shower for at least 1-hour post immersion 5/7 for 2 weeks (10 baths).  IG3 - 60 g/L was filled with 218 L of mineral and 181 L of pure tap water (additionally warmed in special heaters). Bath for 20minutes, soft towel dry and no shower for at least 1-hour post immersion 5/7 for 2 weeks (10 baths). | Passive therapy |
| Ratanasiripong | 2023 | E-wellness programme; 3 x 90min lifestyle and stress management workshops that were also recorded and accessible via YouTube. Online social support via a smartphone app, led an monitored by experience peer leaders | Educational; Psychological |
| Rispinto | 2018 | 12 week multidisciplinary low back pain rehabilitation program (Back on Trek) - teaching self-management of pain by utilising CBT for pain and psychologically informed physical therapy. | Educational; Physical activity; Psychological |
| Rivas | 2018 | dry needle therapy, 18 FM tender-points for 6 weeks | Passive therapy |
| Rivas Neira | 2024 | Aquatic therapy (AT) 3 times weekly for 12 weeks in groups of ten. 60 minutes total, 15 min warm up, 25 mins proprioception, 8 mins stretching, 12 mins relaxation. Clear objectives and progression in intensity, exercises modified to prevent exertion beyond 5 on the Borg scale. Intervention planned for spring to minimise impact of weather on FM symptom severity. Water temperature 30C and depth 120cm  Land based therapy (LBT) 3 times weekly for 12 weeks in groups of ten. 60 minutes total, 15 min warm up, 25 mins proprioception, 8 mins stretching, 12 mins relaxation. Clear objectives and progression in intensity, exercises modified to prevent exertion beyond 5 on the Borg scale. Intervention planned for spring to minimise impact of weather on FM symptom severity. | Physical activity |
| Romero-Zurita | 2012 | Patients participated in three 60-minute Tai-Chi sessions conducted weekly for 28 weeks. Each session included: 15 minutes of warmup with stretching, mobility, and breathing techniques; 30 minutes of Tai-Chi exercises principles and techniques, and finally, 15 minutes of various relaxation methods. Minor modifications were made to adapt for FM | Physical activity |
| Rooks | 2007 | Aerobic and flexibility exercise (AE) - 60 minutes activity, twice weekly. Beginning with 5 mins walking on a treadmill and progressively increasing up to 45 mins walking. Both groups finished with flexibility exercises  Strength training (ST) - 60 minutes activity, twice weekly. Beginning with 5 mins walking on a treadmill and progressively increasing up to 20 mins walking followed by 25 minutes strength training exercises. Both groups finished with flexibility exercises  Fibromyalgia self help course (FSHC) - The FSHC is a 7-session program that teaches individuals with fibromyalgia about the condition and self-management skills. Materials promoted basic self-management techniques to accomplish daily activities and manage symptoms and suggested ways to incorporate wellness activities, including exercise, into daily life. Information was provided through a series of lectures (5-15 minutes) with facilitated group discussion and supplementary readings. Sessions were 120 minutes long every 2 weeks  Strength training and fibromyalgia self help course (ST-FSHC) - participated in ST and FSHC groups | Educational; Physical activity |
| Salas | 2019 | Multidimensional treatment (MDT): individualized exercise program addressing specific physical impairments, as identified by the PI of this study. exercises focused on muscle flexibility and joint range of motion, muscle strengthening, and postural control. twice a week on site with the PI, and to continue the target-specific exercise regimen on their own 1 day per week  Traditional senior exercise therapy (TET)group exercise classes. The exercises included both seated and standing activities focused on improving strength, flexibility, balance, and cardiovascular health. Class exercises were inclusive of daily-living tasks, such as: ascending and descending stairs; chair stands; chair- supported squats; and seated aerobics. Classes were offered three times a week | Physical activity |
| Salokhiddinov | 2020 | Education program - 4 daily classes of 90 minutes | Educational |
| Samhan | 2020 | Group 1: Aquatic exercises (AQBE), water at 32C. Height of water adjusted according to support requirements of each child. 45 minutes including 10 min warm up, 30 min exercise loading and 5 min cool down. Twice per week for 4 weeks, followed by a one  Group 2: Land based exercises (LBE). 10min warm up, 15 minutes aerobic exercise, 15 mins strength training and 5 min cool down stretches. 45 mins in total, twice weekly for 4 weeks, followed by a one month washout period and then 4 weeks of AQBE. | Physical activity |
| San Mauro Martin | 2019 | 500mg turmeric supplement for one month, plus IGUBAC (inflammatory gut brain axis control) diet - low FODMAP, gluten free, low histamine, preservative free and natural food | Nutritional |
| Saral | 2016 | Long term interdisciplinary treatment group (LG) - 10 session extended CBT program (one 3h session per week for 10 weeks), one full day of exercise training and one full day of education  Short term interdisciplinary treatment group (SG) - educational items, exercise training and a brief CBT program over 2 consecutive days | Educational; Physical activity; Psychological |
| Sari | 2023 | Telerehabilitation group - clinical Pilates based exercise program delivered via a YouTube channel, with Zoom meetings and Whatsapp communications to share the exercise links and communicate. Exercises were given twice a day for 8 weeks, sessions approx 10 mins. Weekly video interview via Zoom or Whatsapp, plus an exercise diary. Control group - Exercises in home program taught and given leaflets every 2 weeks with increasingly difficult exercises described. Asked to do exercises twice a day for 8 weeks and keep an exercise diary. Weekly phone calls to assess | Physical activity |
| Sarmento | 2020 | Qigong - "six healing sounds" consisting of three elements; deep diaphragmatic breathing, mild body movements and medication. Participants were trained to practice these elements during 25min practice sessions. Practice twice daily for 10 weeks with a weekly group practice  sham - participants were trained in the same body movements as the Qigong group but the healing sounds combined with diaphragmatic breathing and meditation were not taught | Physical activity; Psychological |
| Scaturro | 2019 | group exercise + laser + TENS | Passive therapy; Physical activity |
| Schiavi | 2013 | Follow a dance routine for 4 months, 1h twice a week. | Physical activity |
| Schweiger | 2020 | Group1 - Migratens nutraceutical supplement (coenzyme Q10, vitamin D, alpha-lipoic acid, magnesium, tryptophan)- 1 sachet twice daily for 12 weeks.  Group 2 - Acupuncture according to principles of traditional chinese medicine. Number and mode of needle insertion personalised based on diagnosis, clinical response and change sin the wrists and tongue. Minimum 30 min session, 2 sessions per week for 10 weeks. | Nutritional; Passive therapy |
| Serrat | 2020 | NAT-FM - active group received 2h sessions consisting of PNE, exercise therapy, CBT, mindfulness training and nature exposure. One session per week for 12 weeks | Educational; Passive therapy; Physical activity; Psychological |
| Serrat | 2021 | FIBROWALK - multicomponenet training based on pain neuroscience education, therapeutic exercise, CBT and mindfulness training. Groups of 20, one 2hr session per week for 12 weeks | Educational; Physical activity; Psychological |
| SevgiUnal Aslan | 2023 | Reiki - six 30min sessions on regions where participants felt pain and other regions corresponding to energy flow  Hand massage - six 30min sessions, 15min for each hand | Passive therapy |
| Sharma | 2022 | Hand reflexology massage, 30min sessions | Passive therapy |
| Sheikh | 2019 | Self-directed Walk With Ease (WWE) program - founded in social cognitive theory conceptual framework. WWE workbook given to all participants to support a 6 week self directed program of walking. | Physical activity |
| Silva | 2022 | During the first month of intervention, a low FODMAPs diet criteria have been added to the anti-inflammatory diet, with the exclusion of foods rich in sugars more fermentable by bacteria. After the first month of intervention, all fruit and vegetables previously excluded were reintroduced, keeping the anti-inflammatory diet for another 2 months, completing a total of 3 months of intervention. | Nutritional |
| Skeates | 2020 | 10 week progressive resistance training (PRT) programme, 1hr supervised session weekly for 10 weeks. | Physical activity |
| Soulat | 2023 | online patient support program delivered over 16 weeks, including physiotherapy sessions, CBT, symptom tracking, lifestyle counselling, disease awareness and self management training | Educational; Passive therapy; Physical activity; Psychological |
| Sousa Fonseca | 2021 | Health education programme 1/7, for 6 weeks. break for a week and then 1 week more. According to the original protocol of the Inter-relational School of Fibromyalgia (ISF), this pause aims at encouraging the autonomy and independence of the participants. meetings lasted 60 minutes  Aquatic Physiotherapy, 1/7 60 minute session at moderate intensity. 1)warming up (5 min); 2) muscle stretching (15 min); 3)free active exercises (30 min); and 4) relaxation (10 min). | Educational; Physical activity |
| Spiritovic | 2021 | 24 week intervention - activity of daily living, resistance and strength (ADLRS) exercises. Supervised 1hr session twice weekly plus 30min home based exercise on remaining 5 days of the week. | Physical activity |
| Spiritovic | 2018 | education material for home exercise plus physical-occupational intervention | Physical activity |
| Stephens | 2008 | Qigong group - 12 week exercise program, 3 sessions per week. 30 minutes supervised 1/7 and then 2/7 unsupervised at home  Aerobics group - 12 week exercise program, 3 sessions per week. 30 minutes supervised, 1/7 with 2/7 at home. | Physical activity |
| Strombec | 2007 | Nordic Walking (walking with specially constructed ski poles). 1 supervised group session per week for 12 weeks, 45 minutes. told to do 45 minutes 2/7 at home.  Range of motion exercises to be performed at home three times a week over 12 weeks. Otherwise, the patients were told to maintain their current level of activity during the 12-week programme. The patients kept exercise logs. | Physical activity |
| Sule | 2019 | Group 1 - Slow-speed resistance training with individualised instruction, targeting the largest muscle groups (leg press, chest press, compound row) performed 1-2 days for 12 weeks.  Group 2 - Instructions to perform aerobic exercise (i.e. walking) for 30 mins, three times weekly, given to participants and their parents. Monthly calls to answer questions. | Physical activity |
| Sveaas | 2017 | 3 month intervention. Two supervised sessions per week performing High Intensity Interval training (HIIT) on treadmill - 4minute walk/run then rest for 3 minutes, repeated 4 times. Followed by 20 minutes of strength for major muscle groups. One individual cardio only session, 40 minutes weekly. | Physical activity |
| Sveaas | 2020 | Two supervised sessions per week for 12 weeks, consisting of high intensity cardiorespiratory and strength exercises. One additional individual cardio exercise session per week | Physical activity |
| Tee | 2016 | Home-based exercise, 2/7 for 4 weeks vs Centre based physical therapy, twice weekly for 4 weeks | Physical activity |
| Thomsen | 2019 | 11 week intervention. The exercise intervention was performed three times per week; two supervised HIIT workouts on a stationary bicycle in groups of max 6, and one additional self-guided HIIT session (e.g. running, cycling, walking uphill). Sessions start with a 10-minute warm-up period followed by 4 Ã— 4 minutes of exercise at 85â€“95% of HRmax interrupted by 3 minutes of exercise at 70% of the HRmax. Encouraged to continue HIIT exercise without guidance during follow-up period (6 months) | Physical activity |
| Thomsen | 2020 | 3 x individual motivational counselling sessions and 0-5 texts each week. The aim was to increase light intensity physical activity and based on behavioural choice theory, constructs of social cognitive theory and motivational interviewing techniques. | Psychological |
| Tiffreau | 2017 | Rehab programme: one month personalised programme focussed on muscle strength, chest expansion, increased ROM, better gait and transfers, increased aerobic capacity. Participants performed 30 minutes on a cycle ergometer three times per week and received three 30 minute massage and relaxation sessions per week. After one month they were given home based exercises based on the hospital protocol | Physical activity |
| Toprak Celanay | 2020 | Intervetnion group - SSE plus kinesiotherapy tape. Applied to stabilise the main muscles of the back  Control group: Spinal stabilisation exercise programme (SSE). The SSE program was carried out 2 days a week for 6 weeks (12 sessions) Each session lasted  about 60 min. It was composed of 10-min warm-up exercises, 40-min  SSE, and 10-min cool-down | Passive therapy; Physical activity |
| Toprak Celenay | 2017 | Exercise group - supervised 1 hour sessions 2 days per week for 6 weeks. 10 min warm up, 40 min aerobic and strengthening exercises, 10 min cool down/stretching  Exercise plus connective tissue massage (CTM) group - as exercise group plus CTM twice weekly for a total of 12 sessions, lasting 5-20 mins each. | Passive therapy |
| Toprak Celenay | 2020 | Group 1 - Exercise program was carried out 2 days a week for 6 weeks under the supervision of a physiotherapist .The fascial correction technique performed with approximately 25% to 50% of tension and oscillated in longitudinal direction on overall back was used  Group 2 - Exercise program  was carried out 2 days a week for 6 weeks under the supervision of a physiotherapist. The Fan Cut kinesio tape technique performed  with 0% tension on the overall back was used | Passive therapy |
| Tore | 2023 | The treatment was the same for both groups and included knee extension, knee flexion, and hip flexion in sitting position; chair sit-up; isometric contraction of quadriceps and hip adductor muscles and straight leg raise in supine position; hip abduction and adduction in the side-lying position; narrow and wide angle squats and standing on one leg in the standing position  Intervetnion group - Supervised telerehabilitation via zoom video conference.24 sessions in total, 3 days a week, for 45-60 min a day and for 8 weeks.  Control group - Given a brochure showing each exercise and explaining how to perform it. Used zoom for assessment points only, exercises performed on their own | Physical activity |
| Torell | 2023 | High Intensity interval training group (HG) - 3 x HIT/7, including two interval training sessions (4x4min) in self-selected activities. They were individually coached and had regular support primarily by digital coaching. | Physical activity |
| Turan | 2014 | IG - Magnotherapy: hip region 20 minutes, During the study, both groups performed range of motion exercises of the hip to enhance better movement patterns, each exercise was given 10 times a day during the period of the study. 15 sessions, one per day. Follow up at 24 weeks  CG - Placebo stimulus applied to the same regions for the same periods as in the magnetotherapy group.  Patients in each group were given a heat pack (20 min) and short-wave treatments applied to bilateral hip region. Both groups performed range of motion  exercises of the hip to enhance better movement patterns, each exercise  was given 10 times a day during the period of the study. Treatment protocol was continued for 1 session per day for 15 days, with follow up at 24 weeks | Passive therapy |
| Turcotte | 2023 | Community based multidisciplinary group intervention - three cohorts of ten individuals. 10 week intervention including a weekly education session and twice weekly group exercise session. Exercise sessions were 1hr and were personalised combinations of aerobic, strength and flexibility exercises. Education sessions were 1h and included 4xCBT strategies, 2 x pain theory and pain care, 2x nutrition and 2 x peer support sessions. All participants were offered referral for consultation at a sleep clinic. Monthly teleconference "team huddles" were held by the healthcare providers to ensure integration of care.  Usual care, 1 hour 1:1 consult with Rheumatologist. | Educational; Nutritional; Physical activity; Psychological |
| Ugurlu | 2017 | Real acupuncture 3 sessions in the first week, 2 sessions/week in the following 2 weeks and 1 session/week in the following 5 weeks (totally 12 sessions) lasting for 30 minutes each session, always performed by the same experienced physician.  Sham acupuncture 3 sessions in the first week, 2 sessions/week in the following 2 weeks and 1 session/week in the following 5 weeks (totally 12 sessions) lasting for 30 minutes each session, always performed by the same experienced physician. | Passive therapy |
| Uhlig | 2016 | Hospital in-patient MDT rehab-focussed programme | Educational; Physical activity; Psychological |
| Valkeinen | 2008 | Concurrent strength and endurance training. 3 training sessions per week for 21 weeks. All strength training sessions were supervised and lasted 60-90 mins. One endurance training session per week was supervised, on alternate weeks when there were two endurance sessions the second was unsupervised. All sessions were performed at a specified level of intensity | Physical activity |
| van Ittersum | 2011 | Booklet about pain neurophysiological mechanisms and central sensitization. The booklet concludes with some case reports of FM patients as examples of how to use the bookletâ€™s information in daily situations, and with an example of a GP explaining the central sensitization mechanism and giving ADL advice to a patient with FM. | Educational |
| van Koulil | 2010 | WLC - two separate groups allocated to either pain avoidance or pain persistence treatment based on screening instrument and therapist judgement  Pain avoidance treatment - 16 twice weekly sessions plus booster session after 3 months. Regular sessions included 2h CBT followed by 2h exercise training. Partners/significant others were invited to attend the 3rd, 9th and 15th sessions. PAT specifically intended to increase daily activity using individual goals and exposure to fear-related situations.  Pain persistence treatment - 16 twice weekly sessions plus booster session after 3 months. Regular sessions included 2h CBT followed by 2h exercise training. Partners/significant others were invited to attend the 3rd, 9th and 15th sessions. Focus on regulating and diminishing PP behaviours using pacing and gradual increase in activity. Realistic goal setting and cognitive restructuring | Physical activity; Psychological |
| van Wilgen | 2007 | Multidisciplinary program - included an education program with seven sessions aiming at: Cognitive restructuring, information on fibromyalgia, goal setting, pacing, distraction, assertiveness training, relaxation and 25 physical therapy sessions aiming at exercises, relaxation and goal setting. The exercises were performed according to the operant conditioning principles using a graded activity program. | Educational; Physical activity |
| Vassalli | 2018 | Progressive resistance training, 2/7 for 12/52. All majr muscle groups. Education programme 1/7 for 5/52  Education only programme - one 1h class per week for 5 weeks | Physical activity |
| Vermaak | 2015 | participants attended 1 session each week for 6 consecutive weeks, with each 2.5 h in duration. Programme focussed on disease specific education topics. Personal development focused on individual weekly goal setting, problem solving and relaxation techniques | Educational; Nutritional; Physical activity; Psychological |
| Vincent | 2013 | Brief multidisciplinary treatment program - 1 week programme. Initial assessment with nurse and internist followed by meeting with exercise and endurance coach to establish personal goals for health and exercise. 3 days of CBT, 6 hours per day. Final visit with internist to plan medication and self management strategies. Family were encouraged to attend all sessions as well as a specific session for them on how to support improved function. Binder of resources and program materials given. Three follow up phone calls at 2 weeks, 1 month and 3 months | Educational; Physical activity |
| Wadell | 2021 | ADIRA trial - crossover trial. 10 weeks of allocated diet, 4mo washout period, 10 weeks of alternative diet regimen. Food ws delivered weekly to all participants at home.  Participants received breakfast composed of orange juice and a mix of yoghurt and quark served with cornflakes or white bread with butter and cheese. The main meals were composed of meat or chicken, potatoes or white rice and high fat dairy. Quark, protein pudding and protein bars were given as snacks. In total, participants were instructed to consume meat ï¿½5 times/wk, limit seafood intake to ï¿½1 time/wk and fruit and vegetables to ï¿½5 portions/day, to choose high fat dairy, use butter as cooking fat and to abstain from probiotics.  Anti-inflammatory diet - Briefly, breakfast contained whole grain, low fat dairy, fruits or berries, nuts (mainly walnuts) and a juice shot containing probiotics (Lactobacillus plantarum v299). The main meals were composed of fatty fish or legumes, potatoes or grains (predominantly whole grain), vegetables and low fat dairy. As snacks, participants received two fruits per day. In total, i.e. including meals not provided by the study, participants were instructed to limit meat consumption to ï¿½3 times/wk, consume ï¿½5 portions/d of fruit, berries and vegetables, to choose low fat dairy and whole grain, as well as to use oil or margarine as cooking fat | Nutritional |
| Walrabenstein | 2023 | Plants for Joints multidisciplinary lifestyle program. Initial meeting with dietician and physical therapist. Groups of 6-12 met 10 times for 2-3h. The group received theoretical and practical education about a whole food plant-based diet (including a cooking class), physical activity and exercise, and stress management. Peer education and support were actively encouraged. Facilitated with videos, exercises for home, detailed weekly menus and daily supplementation with methylcobalamin and cholecalciferol | Educational; Nutritional; Physical activity; Psychological |
| Wang | 2022 | TAU plus 16 week combined exercise program - in person counselling, supervised training, aerobic and functional home based exercises. Exercise intensity was monitored using a heart rate monitor wristband that was synchronised with a smartphone app. Participants were prescribed 30 minutes of moderate intensity exercise, 5 days a week. Their exercise and heart rate data was uploaded via the app. | Educational; Physical activity |
| Wang | 2018 | Clinical nurse, 30 minute appointments, took a history, performed a physical examination, evaluated disease severity, prescribed or recommended medication and dosage changes, monitored the effects of prescribed antiâ€RA medications, educated their patients, instructed them in correct usage of prescribed medication, and provided psychosocial support. The clinical nurse specialist was also involved in ordering blood tests or Xâ€ray  Rheumatologist care as usual | Educational; Psychological |
| Ward | 2018 | Yoga: Group practice consisted of onceâ€weekly 75â€min yoga classes, conducted by a qualified yoga instructor and class assistant. Each class consisted of a 5â€min check in, 5â€min introduction to the class lesson and yoga philosophy theme, 7â€min centring and breathing practice; 10â€min warmâ€up practice, 28â€min session of supine, seated and standing yoga postures (5 âˆ’ 10 postures per class); 15â€min guided relaxation and 5â€min closing discussion.  Home practice consisted of a 20â€min guided relaxation, based on the relaxation technique practised in the group sessions. A CD, recorded by the yoga instructor, was provided. Participants were asked to practise three times per week, at a time and day of their choice  Participants in the usual care group continued with their usual medical care provided by their rheumatologist and general practitioner for the management of their RA | Physical activity |
| Wiliams | 2023 | Mentored group - modelling and reinforcement of disease self-management skills by peers (mentors) to other African American women with SLE (mentees) was achieved through a combination of educational and informal phone or video interactions with each other.  The control condition included small support groups that met on the same schedule as peer mentoring sessions. | Educational |
| Williams | 2010 | Standard care plus Web-enhanced behavioural self-management (WEB-SM). Study participants assigned to the WEB-SM arm were encouraged to explore all aspects of the website and to select those modules that seemed to be of most relevance to their current situation. Participants were encouraged to practice and apply the behavioural skills on a regular basis and to revisit the site when new issues arose. | Psychological |
| Williams | 2014 | Six weekly sessions of the â€œBetter Choice, Better Healthâ€ Chronic Disease  Self-Management Program (CDSMP). About 2 hours each. Topics covered 1) techniques to deal with problems such as frustration, fatigue, pain and isolation, 2) appropriate exercise for maintaining and improving strength, flexibility, and endurance, 3) appropriate use of medications, 4) communicating effectively with family, friends, and health professionals, 5) nutrition, and, 6) how to evaluate new treatments. Classes were highly participative group sessions.  Control group patients received their usual care with Standard care plus Web-enhanced behavioural self-management (WEB-SM). Study participants assigned to the WEB-SM arm were encouraged to explore all aspects of the website and to select those modules that seemed to be of most relevance to their current situation. Participants were encouraged to practice and apply the behavioural skills on a regular basis and to revisit the site when new issues arose.iled book that provided tips for living  a healthy life with a chronic condition | Educational |
| Wong | 2018 | Us12 weeks Tai Chi, 55 minutes, 3/7.  In the first session, the instructor explained the theory behind TC and its procedures providing participants with printed materials on its principles and techniques. In subsequent sessions, participants practiced, under supervised instruction, 10 forms from the classic Yang style of TC.ual lifestyle habits | Physical activity |
| Wu | 2019 | Physical activity counselling: Patients in the intervention group participated in an exercise counselling programme and wore the pedometer on the waist for 1 week to record baseline data and then for 12 weeks. Each of the intervention group participants was taught how to set up the pedometer by the physical activity counsellor. The faceâ€toâ€face counselling was conducted at weeks 1, 4, and 8 at the participants' home or at the clinical office and lasted approximately 30 minutes. Followâ€up phone calls were made on weeks 2, 6, and 10 to assess the participants' achievement of their daily goals, and possible barriers were discussed.  Usual care: patients in the usual care group wore the pedometer for 1 week to register baseline data and then for 12 weeks following the baseline. | Educational; Psychological |
| Xie | 2019 | Exercise group - 12 week 2-phase Baduanjin qijong training program. Phase one: two classes per week for 4 weeks, step by step instructions given and movements adapted to individuals needs, encouraged to practice at home with a DVD. Phase two: home practice at least 3 times weekly, random telephone contacts to encourage and supervise regular performance.  Control - maintain current lifestyle and medication for 12 weeks | Physical activity |
| Yakut | 2021 | Supervised exercise group: combined exercise training consisting of  breathing, resistance and aerobic exercises under the supervision of a physiotherapist twice per week for 12 weeks (in total 24 sessions).  Each session lasted about 1 hour, Thoracic expansion exercises and diaphragmatic breathing exercises were performed together with pursed-lip breathing. Also, at the end of inspiration, exercise of holding at maximum inspiration for 3 seconds was demonstrated to the patients.  Resistance training included exercises for the main muscle groups.  Body weight, free weights and elastic bands were used for resistance training exercises. In SSc patients with pulmonary arterial hypertension (PAH), especially the intensity of upper extremity training, was kept lower  Home exercise group: The patients in the HEG were instructed, in a session of about 1 hour, how to perform the exercise program at home by a physiotherapist. Then, they practiced the exercise program twice per week for 12 weeks at home. The HEG received the combined exercise training consisting of breathing and posture exercise and walking. The warm-up, cool-down and breathing exercises and their contents were the same with the SEG. The patients were contacted by phone or email every 2 weeks | Physical activity |
| Yigit | 2013 | Training given on pathogenesis, clinical manifestations and course of AS as well as preventative measures. Information about physiotherapy was given. Al participants were given home-based exercises including strength and stretching for all major joints and muscles. They were given a manual and a CD to support this and asked to perform the exercises for 30 mins, 5 times a week for 10 weeks.  Half of the participants who were given the training and information for the intervention did not regularly complete the home based exercises the prescribed 30 min 5x weekly. They were therefore treated as a control group. | Physical activity |
| Yip | 2008 | Arthritis Self Management Program (ASMP) - six 2h classes held weekly, 10-15 participants per class. Individuals wrote action plans with the tutor for the stretching, walking and Tai Chi exercises, which were reinforced at each session. Stretching exercises were practices twice as a group a each session and Tai Chi exercises were taught and practiced for 30 mins. | Educational; Physical activity |
| Yoo | 2022 | Progressive muscle relaxation therapy twice a week for 8 weeks  non-PMRT group received conventional physical therapies such as electro-stimulation and heat therapies during this period. | Psychological |
| Yousefi | 2023 | Cognitive-behavioural therapy training was presented to them in groups, by the researcher. The treatment was based on therapeutic packages in 10 sessions of 90 min to change the mental, emotional and behavioural patterns of members  The MBSR treatment program used in this study consisted of 8 sessions of 90 min, once a week as a group, each session began with an exercise and continued with a discussion about the exercise, and ended with homework. | Psychological |
| Youssef | 2021 | Group 1 - isolated upper and lower extremity joint ranges of motion as well as limb movement patterns plus isotonic strength training with low weight loads - 50 minutes, 3/7 for 12 weeks  Group 2 - isolated upper and lower extremity joint ranges of motion as well as limb movement patterns - 50 minutes 3/7 12 weeks | Physical activity |
| Yuen | 2011 | Wii Fit sessions 3/7 for 10 weeks 5-min warm up (e.g. one of the games in Yoga involves stretching and/or deep breathing), after which the participant engaged in the aerobic exer- cise. This was followed by strength training. Each session ended with a 5-min cool-down involving deep breathing and/or slow stepping and walking. | Physical activity |
| Zahiri | 2022 | FAME (focuses on the management of fatigue, pain, nutrition, stress, exercise, and joint protection) - combination of lectures, group discussions, and question and answering methods. | Educational |
| Zangi | 2020 | Vitality Training programme 10 group sessions and 12 weeks physical activity  3-hour group-based education with TAU | Educational; Physical activity |
| Zangi | 2011 | Vitality Training Programme (VTP) - ten 4.5h group sessions over 15 week plus booster session at 6 months. 8-12 participants per group. Each session addressed a specific topic and included mindfulness based exercises, creative exercises and reflection. Between sessions the participants performed awareness and relaxation training by listening to a CD with mindfulness-based exercises and wrote reflective diaries. Facilitators were were specially trained not to give advice or to present  the â€˜right answersâ€™, but to help individuals explore and discover their individual resources and personal values and to find their own meaningful answered  Routine care - offered intervention after completion of study data collection | Psychological |
| Zhou | 2022 | Genuine acupuncture. 4/7 for 4 weeks then 2/7 for 4 weeks. Needles in both groups were in place for 30 minutes  Sham acupuncture using non tapering needles. 4/7 for 4 weeks then 2/7 for 4 weeks | Passive therapy |
