## Appendix 5 for "A scoping review of non-pharmacological interventions for managing fatigue across the lifespan of people living with chronic musculoskeletal conditions"

**Appendix 5. Mechanisms of action to reduce fatigue**

| **Author** | **Year** | **Mechanism** | **MSK condition** | **Detail** |
| --- | --- | --- | --- | --- |
| Abou-Raya | 2014 | Biochemical | PsA | Reduction of inflammation, reduction of obesity-associated inflammation |
| Alentron- Geli | 2008 | Physical, biochemical | Fibromyalgia | Increased strength, increased growth hormone production and cell generation, regulation of metabolic activity |
| Aree-Ue | 2022 | Psychological | OA | Positive health management behaviours |
| Avaux | 2016 | Psychological | SLE | Increased knowledge, validation |
| Azeez | 2020 | Psychological | RA | Positive health management behaviours |
| Baelz | 2023 | Biochemical | Fibromyalgia | Increased neurotransmitter activity |
| Baldi | 2022 | Biochemical | Fibromyalgia | Reduction of oxidative stress, reduction of inflammation |
| Bestas | 2022 | Physical | AxSpA | Muscle relaxation, pain reduction, improved sleep quality |
| Bettoni | 2013 | Biochemical | Fibromyalgia | Reduction of inflammation |
| Bjersing | 2017 | Biochemical | Fibromyalgia | Regulation of metabolic activity |
| Carandang | 2022 | Psychological | scleroderma | Positive health management behaviours, increased knowledge, validation |
| Carbonell Baeza | 2011 | Psychological | Fibromyalgia | Positive health management behaviours |
| Casanueva | 2014 | Biochemical | Fibromyalgia | Reduction of inflammation |
| Cash | 2015 | Physical, biochemical | Fibromyalgia | Improved sleep quality, regulation of cortisol levels |
| Cetin | 2020 | Physical | Scleroderma | Increased aerobic capacity, increased strength |
| Christie | 2015 | Physical, psychological | IRD | Increased aerobic capacity, increased strength, peer support |
| Correia Moretti | 2016 | Biochemical | Fibromyalgia | Increased neurotransmitter activity |
| Davies | 2012 | Physical | SLE | Reduced BMI |
| Diaz-Toral | 2017 | Biochemical | Fibromyalgia | Increased neurotransmitter activity |
| Dover | 2021 | Physical, biochemical | Myositis | Increased strength, regulation of metabolic activity |
| Dures | 2022 | Psychological | IA | Positive health management behaviours, increased self-efficacy |
| Dures | 2012 | Psychological | RA | Validation, increased self-efficacy |
| Elnaggar | 2022 | Physical, biochemical | JIA | Muscle relaxation, pain reduction, improved microcirculation, reduction of inflammation |
| Elnaggar | 2021 | Physical, psychological | Myositis | Increased strength, peer support |
| feldthusen | 2016 | Psychological | RA | Positive health management behaviours |
| Fonseca | 2021 | Biochemical | Fibromyalgia | Regulation of cortisol levels, reduction of inflammation |
| Gavilan Carrera | 2022 | Physical | SLE | Increased aerobic capacity, increased strength |
| Givilan Carrera | 2023 | Physical, biochemical | Fibromyalgia | Increased strength, regulation of metabolic activity |
| Gizinska | 2015 | Biochemical | RA | Stimulation of the endocrine system, regulation of cortisol levels, regulation of the sympathetic nervous system |
| Gok Metin | 2016 | Physical | RA | Pain reduction |
| Groessl | 2020 | Physical | CLBP | Pain reduction |
| Gyulai | 2015 | Biochemical | MSK disease | Improved microcirculation |
| Hakimi | 2020 | Physical, psychological | EDS | Increased strength, fear reduction |
| Hammond | 2008 | Psychological | Arthritis | Positive health management behaviours |
| Harper | 2021 | Psychological | Vasculitis | Increased self-efficacy |
| Hasni | 2022 | Biochemical | SLE | Regulation of metabolic activity |
| Hewlett | 2019 | Psychological | Arthritis | Positive health management behaviours |
| Jastrzabek | 2013 | Biochemical | RA | Stimulation of the endocrine system, regulation of the immune system |
| Kabiri | 2018 | Biochemical | OA | Stimulation of the endocrine system |
| Kilic | 2023 | Physical | RA | Muscle relaxation |
| Kim | 2023 | Physical, biochemical | CLBP | Muscle relaxation, pain reduction, increased growth hormone production and cell generation, regulation of metabolic activity, regulation of the immune system |
| Lindqvist | 2017 | Biochemical | RA | Reduction of inflammation |
| Liu | 2012 | Biochemical | Fibromyalgia | Regulation of cortisol levels |
| Maarj | 2023 | Physical | Hypermobility | Increased strength |
| Maddali Bongi | 2016 | Physical | Fibromyalgia | Muscle relaxation |
| Maddali Bongi | 2011 | Physical | Fibromyalgia | Pain reduction |
| Maddali Bongi | 2019 | Psychological | Sjogrens | Reduction of stress |
| McCormack | 2018 | Psychological | IRD | Increased self-efficacy, acceptance |
| Mhalla | 2011 | Biochemical | Fibromyalgia | Regulation of cortisol levels |
| Miyamoto | 2019 | Physical | Sjogrens | Increased aerobic capacity |
| Murphy | 2023 | Psychological | Scleroderma | Peer support, increased self-efficacy |
| Neuburger | 2007 | Physical | RA | Increased aerobic capacity, increased strength |
| O'Connor | 2013 | Physical | Chronic MSK pain | Muscle relaxation, pain reduction |
| Oksuz | 2023 | Physical | AxSpA | Pain reduction |
| Paek | 2016 | Psychological | RA | Positive health management behaviours |
| San Mauro Martin | 2019 | Biochemical | Fibromyalgia | Reduction of oxidative stress, reduction of inflammation |
| Sanchez | 2019 | Biochemical | Fibromyalgia | Improved microcirculation, reduction of inflammation |
| Sarmento | 2020 | Biochemical | Fibromyalgia | Stimulation of the endocrine system, regulation of the immune system |
| Sveaas | 2020 | Physical, biochemical | AxSpA | Pain reduction, increased aerobic capacity, increased strength, reduction of inflammation |
| Sveaas | 2018 | Psychological, biochemical | AxSpA | Peer support , increased self-efficacy, stimulation of the endocrine system, increased neurotransmitter activity, reduction of inflammation |
| Thomsen | 2019 | Physical | PsA | Increased aerobic capacity |
| Toprak Celenay | 2017 | Physical | Fibromyalgia | Increased strength |
| Vermaak | 2015 | Psychological | RA | Fear reduction |
| Wong | 2018 | Physical, biochemical | Fibromyalgia | Pain reduction, increased aerobic capacity, increased neurotransmitter activity |
| Yakut | 2021 | Physical | Scleroderma | Increased aerobic capacity, increased strength |
| Yuen | 2011 | Psychological | SLE | Positive health management behaviours |
| Zangi | 2012 | Psychological | IRD | Reduction of stress |
| Zhou | 2022 | Biochemical | Sjogrens | Regulation of the sympathetic nervous system |
