## Appendix 6 for "A scoping review of non-pharmacological interventions for managing fatigue across the lifespan of people living with chronic musculoskeletal conditions"

Appendix 6: Fatigue measurement tools – summary

| **Fatigue measurement tool** | **Occurrences** | |
| --- | --- | --- |
|  | **Fatigue is main focus of study** | **Total** |
| Bath Ankylosing Spondylitis Disease Arthropathy Index (BASDAI) | **1** | **6** |
| Bath Ankylosing Spondylitis Functional Index (BASFI) | **0** | **1** |
| Cognitive Exercise Therapy Approach – Biopsychosocial Questionnaire (BETY-BQ) | **1** | **1** |
| Brief Fatigue Inventory (BFI) | **3** | **5** |
| Bristol Rheumatoid Arthritis Fatigue (BRAF) Multi-Dimensional Questionnaire | **4** | **8** |
| Chalder Fatigue Scale | **3** | **3** |
| Checklist Individual Strength (CIS) | **1** | **5** |
| EULAR Sjogren’s Syndrome Patient Reported Index (ESSPRI) | **1** | **2** |
| Functional Assessment of Chronic Illness Therapy (FACIT) | **5** | **18** |
| Fatigue Assessment Scale (FAS) | **0** | **2** |
| Fatigue Impact Scale (FIS) | **1** | **5** |
| Fatigue Severity Scale (FSS) | **16** | **43** |
| FibroFatigue Scale (FFS) | **0** | **2** |
| Flinders Fatigue Scale | **0** | **1** |
| Fibromyalgia Impact Questionnaire (FIQ) | **5** | **35** |
| Fibromyalgia symptom form | **0** | **1** |
| Global Fatigue Index (GFI) | **1** | **4** |
| Combined Index of Severity of Fibromyalgia (ICAF) | **1** | **1** |
| Kids Fatigue Severity Scale (K-FSS) | **0** | **1** |
| Likert scale | **0** | **1** |
| Lupus Quality of Life (LupusQoL) | **1** | **4** |
| Multidimensional Assessment of Fatigue (MAF) | **5** | **14** |
| Multidimensional Fatigue Inventory (MFI) | **9** | **26** |
| Multidimensional Fatigue Symptom Inventory – Short Form (MFSI-SF) | **0** | **1** |
| Numerical Rating Scale (NRS) | **3** | **10** |
| Paediatric Quality of Life (PedsQL) | **1** | **8** |
| Pichot Fatigue Scale | **0** | **2** |
| Piper Fatigue Scale | **1** | **2** |
| Profile of Mood States (POMS) - fatigue | **2** | **5** |
| Profile of Fatigue and Discomfort Sicca Symptoms Inventory (PROFAD-SSI) | **1** | **2** |
| Patient-Reported Outcomes Measurement Information System (PROMIS) | **4** | **10** |
| Rheumatoid Arthritis Impact of Disease (RAID) | **0** | **1** |
| Silhouettes Fatigue Scale (SFS) | **0** | **1** |
| Swedish Occupational Fatigue Inventory (SOFI) | **1** | **1** |
| Visual Analogue Scale (VAS) | **17** | **60** |
